# Phenotype Progression of Complex Regional Pain Syndrome Identified by Quantitative Sensory Testing

**DOI:** 10.1101/2025.08.05.25333054

**Authors:** Jonathan R. Husk, David Pang, Fauzia Hasnie, Andreas Goebel, Walter Magerl

## Abstract

Complex Regional Pain Syndrome (CRPS) exhibits persistent disproportionate limb pain and hyperalgesia associated with neuroinflammatory and autonomic changes, typically after inciting limb injury. However, little is known about the progression of somatosensory changes over time. We reviewed cross-sectional studies employing standardised Quantitative Sensory Testing (QST) in accordance with the DFNS comprehensive somatosensory test protocol, stratified for CRPS duration.

This study was registered with PROSPERO ID CRD42020216485. Reporting follows PRISMA guidelines. Databases searched were PubMed, Cochrane Library, Google Scholar, EMBASE, Web of Science and Scopus. Studies of adult patients with CRPS and QST according to the DFNS protocol were included. 1415 articles were screened and 23 studies meeting the inclusion criteria were included in quantitative analysis and narrative synthesis. Analysis was stratified by CRPS duration into an early (≤ 6 months), intermediate (6-12 months) and late (> 12 months) time window.

Across all studies encompassing 2059 CRPS patients all somatosensory parameters deviated significantly from healthy subject profiles (all *P* < 0.001). All non-nociceptive detection parameters displayed a significant loss of sensitivity, while all nociceptive parameters as well as thermal and mechanical dysesthesias, i.e., paradoxical heat sensation and allodynia displayed a significant gain of sensitivity. Pressure pain threshold (PPT) showed the most drastic sensory gain with a large effect size (> 2 SD; Hedges’ *g* = 0.9) equivalent to more than half of CRPS patients exhibiting abnormal pressure hyperalgesia. Moreover, PPT significantly increased progressively with increasing CRPS duration (from 1.7 to 2.2 and 2.8 SD above normal). In late CRPS (> 12 months), additionally *contralateral*, mirror image test site sensory loss (*P* < 0.001) and pressure hyperalgesia (*P* < 0.001) became evident. Stratification for magnitude of pain did only modestly predict relevant differences in somatosensory profiles.

Quantitative analysis of 23 cross-sectional QST studies in CRPS demonstrates pressure hyperalgesia as the ‘signature’ CRPS sensory abnormality, distinguishing CRPS from neuropathic pain conditions. Further, pressure hyperalgesia exhibits a peculiar dramatic step progression between 4-12 months, without equivalent in other parameters. In late CRPS, somatosensory profiles become significantly abnormal at contralateral areas indicating loss of regionality. These findings have important implications for the classification of CRPS as a chronic primary pain condition, for understanding challenges to rehabilitative interventions, and for condition-subtyping.

## Introduction

Complex Regional Pain Syndrome (CRPS) is a rare, severe limb pain condition triggered by, often trivial inciting events resulting in disproportionate clinical signs and symptoms, including sensory, oedema, vasomotor, sudomotor, trophic, and motor abnormalities.^1–5^ The CRPS incidence in the general population is low (< 1/10.000), however, it may be high in at-risk-populations (> 3%), e.g., following limb trauma and/or distal limb surgery.^6,7^ CRPS occurs with a threefold higher preponderance in females and may affect upper limbs more frequently than lower limbs.^3,5^

Persistent severe and debilitating limb pain is the predominant symptom, typically perceived in deep tissue, although occasionally superficial pain can coexist; pain is often extremely exacerbated by joint movement, pressure or even light touch.^1,8–10^ Pain quality varies and is typically described neuropathic-like, such as sharp, burning, tearing and stinging sensations. There is no FDA/EMA-approved treatment, and no high-certainty evidence exists for the effectiveness of any pharmacological intervention treating CRPS.^11,12^

The Budapest criteria with the recent Valencia adaptation remain the most accepted and current means of diagnosing CRPS.^13,14^ It requires assessment of both clinical symptoms and signs while excluding other diagnoses that could better explain these clinical features. Whereas specific investigations such as imaging and neurophysiology can support the diagnosis by excluding other conditions, they are not required to diagnose CRPS.

CRPS is recognised as a *chronic primary pain syndrome* in the recent amendment of ICD-11, denoting ‘pain in its own right’, i.e., not secondary to established disease.^13,15^ The pathophysiology of CRPS remains incompletely understood and likely involves early inflammatory mechanisms and IgM-(early)/IgG-mediated (late) autoimmunity, amongst _others._5,10,16,17

CRPS I and CRPS II subtypes are distinguished by the identified absence or presence of injury to a major nerve, however, are clinically almost identical, and the relevance of this distinction is uncertain.^13^ Other CRPS subclassifications have been proposed related to purported autonomic, peripheral/central nervous system, genetic, inflammatory and autoimmune mechanisms, and involving the time course, i.e., early versus late CRPS^10,18–22^; however the relevance of these groups has remained unclear. Attempts at phenotype subclassification have not resulted in effective management approaches.^14^ About 80% of patients with CRPS naturally improve over the course of the first 12-15 months, whereas any improvement is very rare thereafter.^2,23^

It is unknown whether patients with longstanding CRPS (> 12-15 months) differ in their somatosensory profiles from those with early CRPS, and further whether any such differences might already define patients early in their condition, so that they might support early subtyping. Understanding this would allow enhanced patient information and stratified research involving patients early in their disorder.

Recent guidelines support the use of Quantitative Sensory Testing (QST) in chronic pain; such testing allows for detailed somatosensory profiling of patients in the diagnosis of neuropathic pain.^24–28^ The comprehensive QST protocol proposed by the German Research Network on Neuropathic Pain (DFNS) constitutes the current gold standard in neuropathic pain assessment.^26,27,29^ It allows the identification of patient subgroups in a mechanism-related manner, and it is an approved tool for patient stratification in studies on medicinal products for pain treatment by the European Medicines Agency.^30,31^

Reference data in healthy subjects for different body areas (face, hands, feet and trunk) further stratified for sex and age allow data normalisation independent of these parameters, and direct comparison of somatosensory modalities independent of their physical dimension.^26,27,32^

Previous studies using the DFNS QST protocol discriminated CRPS from neuropathic pain by the widely differing somatosensory profiles.^33,34^ Accordingly, QST may provide insight into pain mechanisms in CRPS.^21,28,35^ Only recently, however, have there been efforts to aggregate the knowledge on CRPS somatosensory profiles.^21,35^ No systematic efforts have been made, to our knowledge, to gain insight into the somatosensory abnormalities across the natural history of CRPS.

In this systematic review and evidence synthesis we strove to take advantage of the high degree of standardisation acquired by different groups using the DFNS protocol for CRPS phenotyping over the preceding two decades; the objective was to identify changes in somatosensory profiles obtained through standardised QST across different time brackets of CRPS duration. We conducted a systematic literature research and included patient data from 23 studies. The population included 2059 patients over 18 years of age with a diagnosis of CRPS according to the Budapest criteria^13^; data was stratified by CRPS duration into an early (< 6 months), intermediate (6-12 months) and late subgroup (> 12 months).

## Materials and methods

This study follows the Preferred Reporting Items for Systematic Reviews and Meta-Analyses (PRISMA) statement^36^ and was preregistered on PROSPERO (CRD42020216485).

### Selection of studies

DP, FH and JRH conducted the literature search, DP and JRH extracted the data, DP performed the risk of bias assessment. In accordance with PRISMA guidelines, an electronic literature search was performed. We systematically reviewed studies of CRPS that included QST following the DFNS protocol. A literature search was performed using these databases:

1. PubMed
2. Cochrane Library
3. Google Scholar
4. EMBASE
5. Web of Science
6. Scopus

Dates searched were from inception to February 28, 2025. After screening of titles and abstracts full texts were retrieved for eligible studies. The search strategy used is shown in Box 1.

#### Box 1

**Search strategy in systematic literature research**

(Complex Regional Pain Syndrome *OR* CRPS *OR* Reflex sympathetic dystrophy *OR* Causalgia *OR* Algodystrophy *OR* Sudeck’s atrophy *OR* Shoulder hand syndrome *OR* Neurodystrophy *OR* Neuroalgodystrophy *OR* Reflex neuromuscular dystrophy *OR* Posttraumatic dystrophy) *AND* (Quantitative sensory testing *OR* QST *OR* Thresholds *OR* sensory *OR* Thermal *OR* Mechanical *OR* Heat *OR* Cold *OR* Hyperalgesia *OR* Allodynia)

Language was restricted to English and only peer-reviewed adult studies included. Reference lists of the papers used were searched manually for relevant articles and potential references. Citing articles were searched for electronically to ensure articles meeting the inclusion criteria were not missed.

### Inclusion/exclusion criteria

Studies included in this systematic review:

1. Subjects investigated had a diagnosis of complex regional pain syndrome or older diagnostic terms for this condition.
2. Measured sensory profiles by QST using mechanical and/or thermal stimuli applied according to the DFNS protocol (supplement 1 provides an overview of QST parameters and their roles in somatosensory perception).
3. QST protocol describes stimulus modality, anatomical site and intensity.
4. Published in a peer-reviewed journal in English.

Studies excluded from this systematic review:

1. Duplicate publications or follow-up analysis of previously published data.
2. Conference abstracts, book chapters or dissertations.
3. Studies involving patients younger than 18 years of age.

### Data extraction

Data were extracted based on the following: definition of CRPS, geographical location, type of CRPS (I or II), disease duration, year of publication, sample size, age distribution, sex (percent female), current/average pain intensity, anatomical site, comparison/control site, QST measures (z-scores) of the DFNS protocol (for details on parameters of the DFNS protocol see supplement 1).

### Data synthesis and statistical analysis

QST profiles were calculated across included studies by calculating the weighted mean for z-scores, and the weighted mean for standard deviation, from which in turn the standard error and 95% confidence interval were calculated. The weighted means were calculated as follows, with w being the weights (sample size of respective study) and x being the z-score of the respective study:

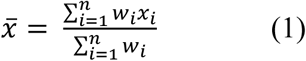

Studies were then stratified by CRPS duration into three subgroups: early (CRPS duration < 6 months), intermediate (6-12 months) and late (> 12 months). We decided on these subgroups since by far the largest group of patients begin to dramatically improve within the first 6 months and conversely patients rarely improve later than 12-15 months after disease onset.^14,23^ For each subgroup weighted z-scores and standard deviation, standard error and 95% confidence interval were calculated. Stratification was done by sorting studies into subgroups according to mean CRPS duration (Table 1). Some of the studies reported median instead of mean and either range or interquartile range of data. In these cases, we estimated mean and standard deviation by using approximating standard transformation equations (see supplement 3). We also stratified studies by mean pain ratings on a numerical rating scale (NRS 0-10) into two subgroups (NRS < 5; NRS ≥ 5) to assess their influence on QST profiles. Groups and subgroups were statistically compared with two-way ANOVA with Bonferroni multiple comparisons in GraphPad Prism 10.4.1. As an estimator for effect size, we calculated Hedges’ g.^37^

**Table 1.** Study and subgroup characteristics for studies included in stratification by CRPS duration.

|  | n | n(female) | %(female) | Age (years) | NRS (0-10) | CRPS duration (months) |
| --- | --- | --- | --- | --- | --- | --- |
| **Early time window (CRPS duration < 6 months)** |  |  |  |  |  |  |
| Wittayer 2018 (Early) | 20 | 15 | 75.0 | 57.7 ± 12.7 | 4.2 ± 2.5 | 1.6 ± 1.0 |
| Louis 2025 | 85 | 64 | 75.3 | 52.9 ± 13.3 | 4.7 ± 2.3 | 2.5 ± 1.5 |
| Mainka 2013 | 18 | 7 | 38.9 | 51.7 ± 10.1 | 5.8 ± 1.8 | 3.3 ± 2.6 |
| Mehling 2024 | 20 | 16 | 80.0 | 49.0 ± 12.0 | 5.4 ± 1.2 | 3.4 ± 4.7 |
| Huge 2008 (Early) | 27 | 25 | 92.6 | 56.0 ± 12.8 | 2.8 ± 2.6 | 3.5 ± 2.1 |
| Üceyler 2007 | 32 | 22 | 68.8 | 52.6 ± 11.8 | 4.8 ± 2.7 | 4.4 ± 3.9 |
| Eberle 2009 | 50 | 36 | 72.0 | 46.0 ± 10.0 | n/a | 5.0 ± 0.7 |
| Kumovski 2017 | 24 | 14 | 58.3 | 51.6 ± 9.8 | 4.7 ± 2.1 | 5.6 ± 2.7 |
| Reimer 2016 | 19 | 15 | 78.9 | 58.1 ± 13.8 | 5.6 ± 3.0 | 5.7 ± 8.3 |
| Enax-Krumova 2017 | 24 | 15 | 62.5 | 50.9 ± 11.3 | 6.0 ± 2.0 | 5.8 ± 3.9 |
|  | **319** | **229** | **71.8 ± 12.0** | **52.1 ± 11.9** | **4.8 ± 2.3** | **3.9 ± 2.6** |
| **Intermediate time window (CRPS duration 6-12 months)** |  |  |  |  |  |  |
| Dietz 2019 | 105 | 86 | 81.9 | 52.8 ± 12.6 | 5.0 ± 2.0 | 7.2 ± 6.9 |
| Habig 2021 | 10 | 6 | 60.0 | 33.0 ± 9.8 | 6.7 ± 1.1 | 9.4 ± 8.7 |
| Dimova 2020 | 202 | 156 | 77.2 | 51.2 ± 12.9 | 4.8 ± 2.3 | 9.6 ± 20.6 |
| König 2019 | 52 | 32 | 61.5 | 50.0 ± 11.4 | 3.9 ± 2.5 | 9.7 ± 6.5 |
| Wittayer 2018 (Late) | 33 | 25 | 75.8 | 52.2 ± 12.8 | 4.4 ± 2.1 | 10.0 ± 8.0 |
| Breuer 2014 | 20 | 10 | 50.0 | 47.8 ± 8.0 | 4.7 ± 2.0 | 11.0 ± 6.0 |
| Maier 2010 | 403 | 312 | 77.4 | 52.0 ± 13.0 | 5.8 ± 2.4 | < 1 year n = 214,<br>> 1 year n = 189<sup>a</sup> |
| Meyer-Frießem 2020 | 339 | 262 | 77.3 | 52.0 ± 13.6 | 4.5 ± 3.2 | < 1 year n = 179,<br>> 1 year n = 157,<br>unknown n = 3<sup>a</sup> |
|  | **1164** | **889** | **76.4 ± 5.7** | **51.6 ± 12.9** | **5.0 ± 2.6** | **11.0 ± 13.5<sup>a</sup>** |
| **Late time window (CRPS duration > 12 months)** |  |  |  |  |  |  |
| Gierthmühlen 2012 | 344 | 271 | 78.8 | 51.6 ± 13.2 | 5.8 ± 2.3 | 21.7 ± 35.2 |
| Schmidt 2025 | 11 | 9 | 81.8 | 50.6 ± 8.0 | 6.8 ± 2.3 | 36.0 ± 39.8 |
| Huge 2008 (Late) | 34 | 29 | 85.3 | 61.6 ± 12.5 | 2.3 ± 2.7 | 37.4 ± 15.1 |
| Huge 2011 | 118 | 91 | 77.1 | 58.0 ± 12.0 | 2.8 ± 2.7 | 42.0 ± 23.0 |
| de Schoenmacker 2023 | 21 | 17 | 81.0 | 44.0 ± 12.0 | 5.5 ± 2.4 | 59.9 ± 38.3 |
| van Rooijen 2013 | 48 | 35 | 72.9 | 46.4 ± 12.1 | 6.2 ± 2.1 | 120.0 ± 88.8 |
|  | **576** | **452** | **78.5 ± 2.7** | **52.8 ± 12.7** | **5.0 ± 2.4** | **36.7 ± 36.2** |
| One-way ANOVA p value |  |  | < 0.0001 | 0.18 | 0.16 |  |
| **Entire population** | **2059** | **1570** | **76.3 ± 6.3** | **52.0 ± 12.7** | **5.0 ± 2.5** | **17.1 ± 18.2** |

Mean and SD have been estimated for Enax-Krumova 2017 (age, NRS, CRPS duration), Breuer 2024 (age, NRS, CRPS duration), de Schoenmacker 2023 (CRPS duration), and Wittayer 2018 (CRPS duration), see methods section for details. All three subgroups were statistically compared with one-way ANOVA for the proportion of females, age and mean pain ratings. Values are displayed as mean ± SD; n/a: data not available; NRS: mean pain rating on a numerical rating scale.
<sup>a</sup>Mean duration was estimated at 12 months for Maier 2010 and Meyer-Frießem 2020 to calculate the weighted mean CRPS duration of the intermediate group as they only reported the number of included patients with a disease duration below and above one year.

## Results

### Literature search

3564 studies were identified from which 1415 articles remained after duplicates were removed. Following screening of titles and abstracts, 48 studies were identified for assessment of the full text. 23 studies with a total of 2059 participants were included in quantitative analysis (Fig. 1). The remaining studies were either not performing QST according to the DFNS protocol or were using data that had been published previously in a different context (see supplement 2 for details).

**Figure 1.**
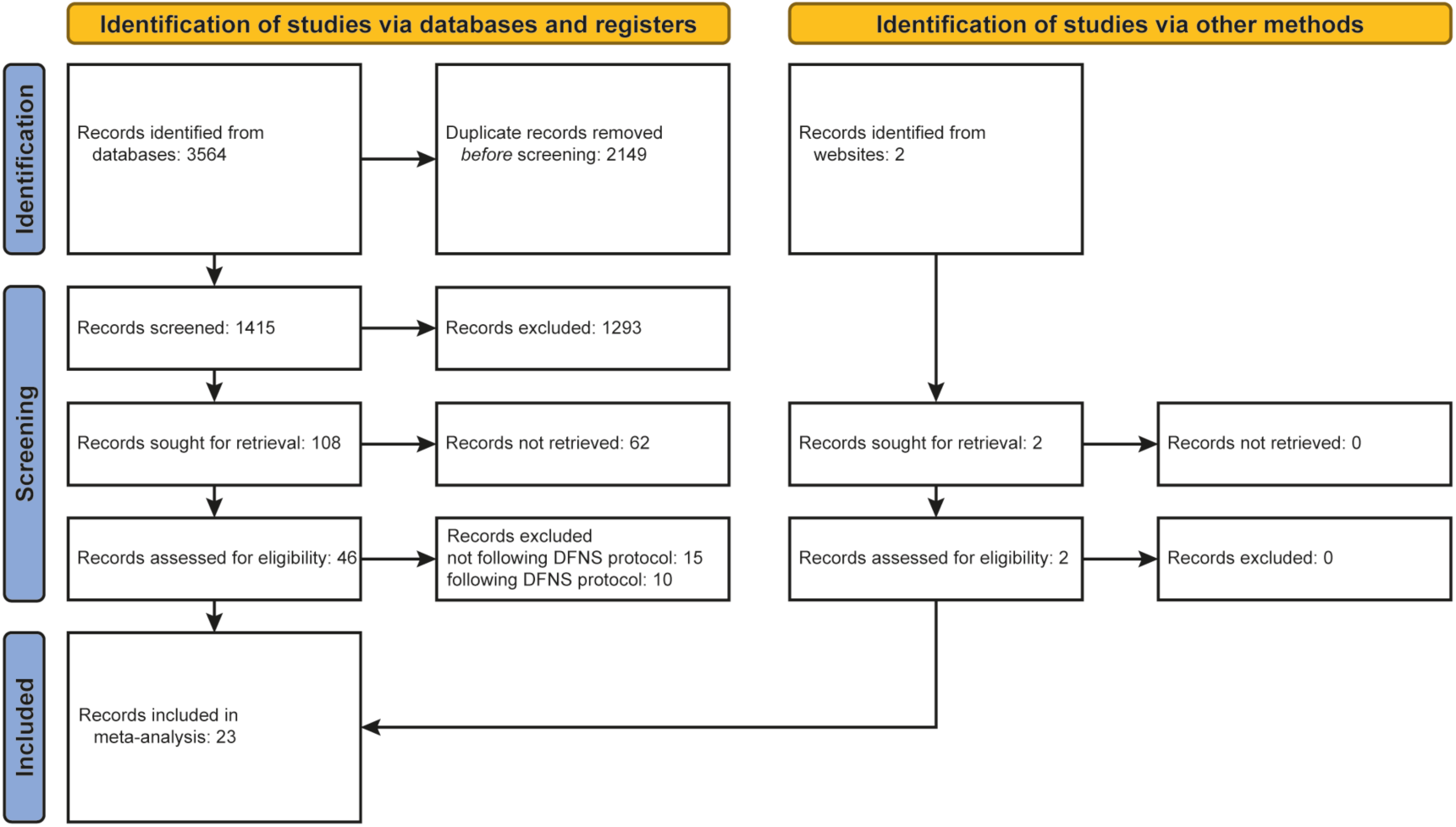
Prisma flow chart of systematic literature research. Studies were excluded if they were not published in a full article format or if data could not be extracted (although attempts were made with the corresponding authors where necessary). Case reports were excluded for the systematic review of QST results; but other specific study designs were not restricted, to maximise retrieval of potentially relevant studies. Mixed adult and paediatric studies were excluded. No distinction between CRPS type I and II was made. All articles were screened by two reviewers independently. Articles that did not meet the inclusion criteria were removed from analysis. For the quantitative analysis, QST profiles from 23 out of 48 studies were included. Supplement 2 provides an overview on all 48 studies with brief statements on reasons for non-inclusion of 25/48 studies.

### Risk of bias assessment

Risk of bias assessment of included studies is presented in supplement 4. All studies reported the research question or objectives clearly [D1]. Only one study did not clearly specify the study population [D2]. Two studies did not specify whether subjects were recruited from the same or similar populations [D3]. One study did not clearly define and implement QST parameters across populations [D4]. Three studies did not provide mechanism-based justification for the use of QST [D5]. All studies clearly defined their outcome measures across study participants [D6].

### Study characteristics

For studies included in quantitative analysis, study and subgroup characteristics are given in Table 1. We stratified the QST profiles by CRPS duration in an early, intermediate, and late group (Table 1). Within the 23 reviewed publications, the early group was derived from 10 cohorts, a total of 319 patients, the intermediate group from 8 cohorts comprising 1164 patients, and the late group from 6 cohorts including 576 patients (Table 1). The total number of cohorts was therefore 24; while patients included in most of the 23 publications were classed by us into one of these three groups (i.e., one group per publication), two publications^38,39^ had reported data already stratified for both acute and chronic subgroups, so that each of these provided two cohorts; whereas one cohort was reported in two publications^40,41^ explaining the overall cohort number.

Contralateral data was reported for 5 cohorts, a total of 201 patients in the early group, by 5 cohorts, 772 patients in the intermediate group, and for 4 cohorts for a total of 221 patients in the late group. For an overview of QST data in the affected and contralateral body side reported in the included studies, see supplement 5.

### Somatosensory profiles in CRPS

Most studies reported the complete QST profile (13 parameters). The remaining studies were missing one or few of the parameters, which did not affect the number of patients to an appreciable extent (< 10% loss of data except for WUR (13%), DMA (21%), and PHS (15%); for details see supplement 2). In eleven studies QST was available for the contralateral mirror image site, in two studies QST was also performed in an additional control site.

All parameters of the QST profile in the area affected by CRPS deviated highly significantly from the profile of healthy controls (all *P* < 0.001, Fig. 2). While there was a general loss in thermal and mechanical sensitivity (increased detection thresholds), there was a general gain in thermal and mechanical pain thresholds, or pain sensation or pain summation (lower thresholds or higher pain ratings). CRPS patients also exhibited thermal (PHS) and mechanical dysesthesia (DMA) although their magnitude was modest (Fig. 2A, supplement 5). Effect sizes were moderate-to-large (0.5 < *g* < 0.8) for all thermal QST parameters (CDT, WDT, TSL, CPT, HPT) and vibration detection (VDT). Effect sizes were small-to-moderate (0.2 < *g* < 0.5) for all punctate mechanical stimuli (MPT, MPS, WUR, MDT) and dysesthesia parameters (DMA, PHS). The only QST parameter exceeding the threshold for a large effect size was pressure pain (PPT; Hedges’ *g* = 0.90; Fig. 2B).

**Figure 2.**
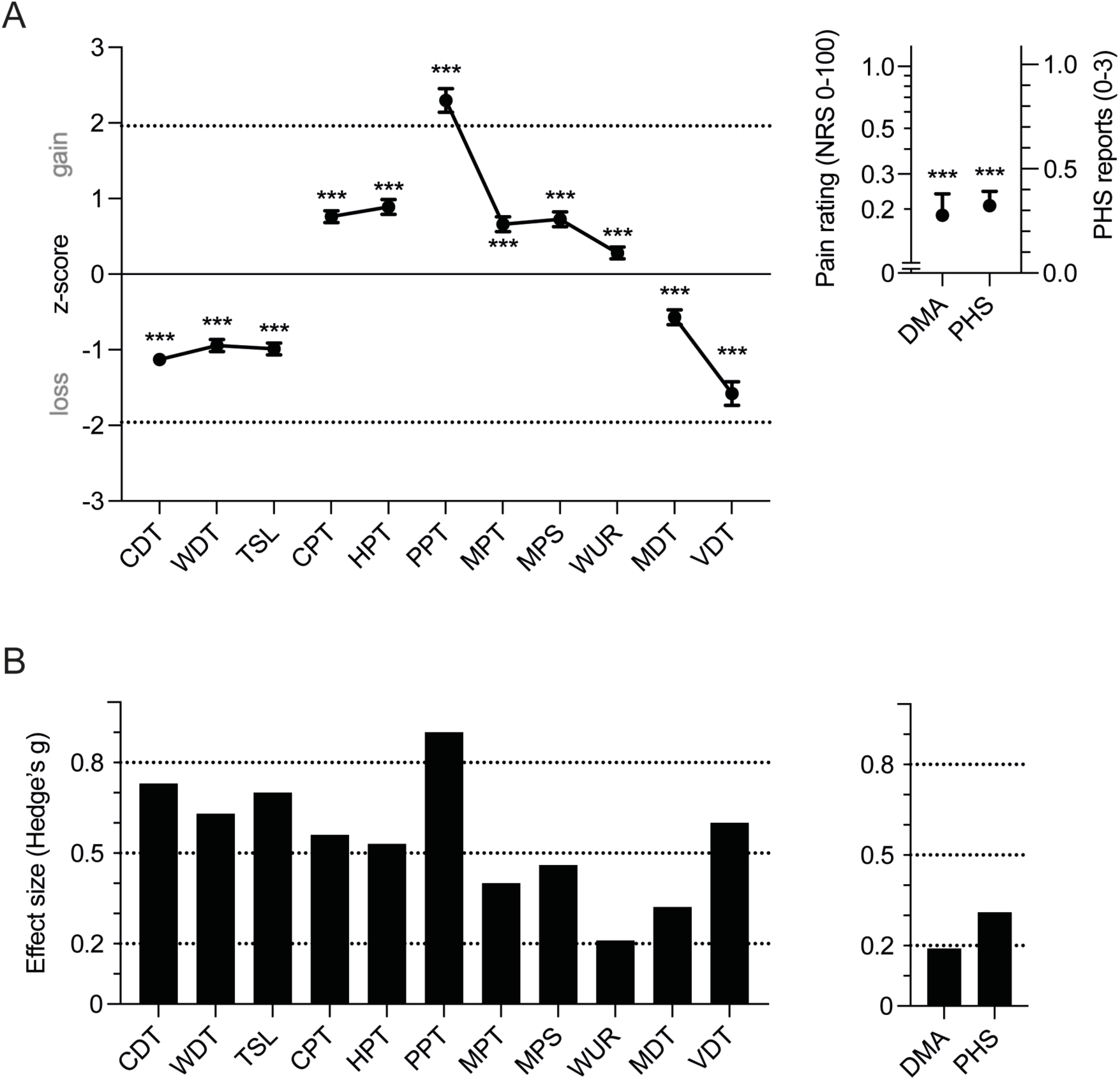
Overall somatosensory profile at the ipsilateral site (affected area) across all studies included in meta-analysis. **(A)** Weighted mean z-scores for all QST parameters across all studies included in meta-analysis (mean ± 95% confidence interval). DMA and PHS are reported as absolute values as z-transformation is not possible.^32^ Dashed lines indicate deviation of 1.96 standard deviations from the mean of the reference population. Asterisks indicate statistical significance compared to the DFNS healthy reference population (*** *P* < 0.001). **(B)** Effect size (Hedges’ g) for the change compared to the healthy DFNS reference population. Dashed lines indicate the distinction between no relevant difference and weak effect size (*g* = 0.2), between weak and moderate effect size (*g* = 0.5), and between moderate and strong effect size (*g* = 0.8). CDT: cold detection threshold, WDT: warm detection threshold, TSL: thermal sensory limen, CPT: cold pain threshold, HPT: heat pain threshold, PPT: pressure pain threshold, MPT: mechanical pain threshold, MPS: mechanical pain sensitivity, WUR: wind-up ratio, MDT: mechanical detection threshold, VDT: vibration detection threshold, DMA: dynamic mechanical allodynia, PHS: paradoxical heat sensations.

A subset of studies reported both ipsi- and contralateral QST. *Ipsilateral* QST in this subset was well representative of the complete data set described above (Fig. 3A). and accordingly, effect sizes were similar as follows: they were for CDT and TSL large (g > 0.8), for WDT and PPT just below the cut-off between a moderate and large effect size (both g = 0.78), and effect sizes for CPT, HPT, MPT, MPS, VDT and DMA were moderate (Fig. 3B).

**Figure 3.**
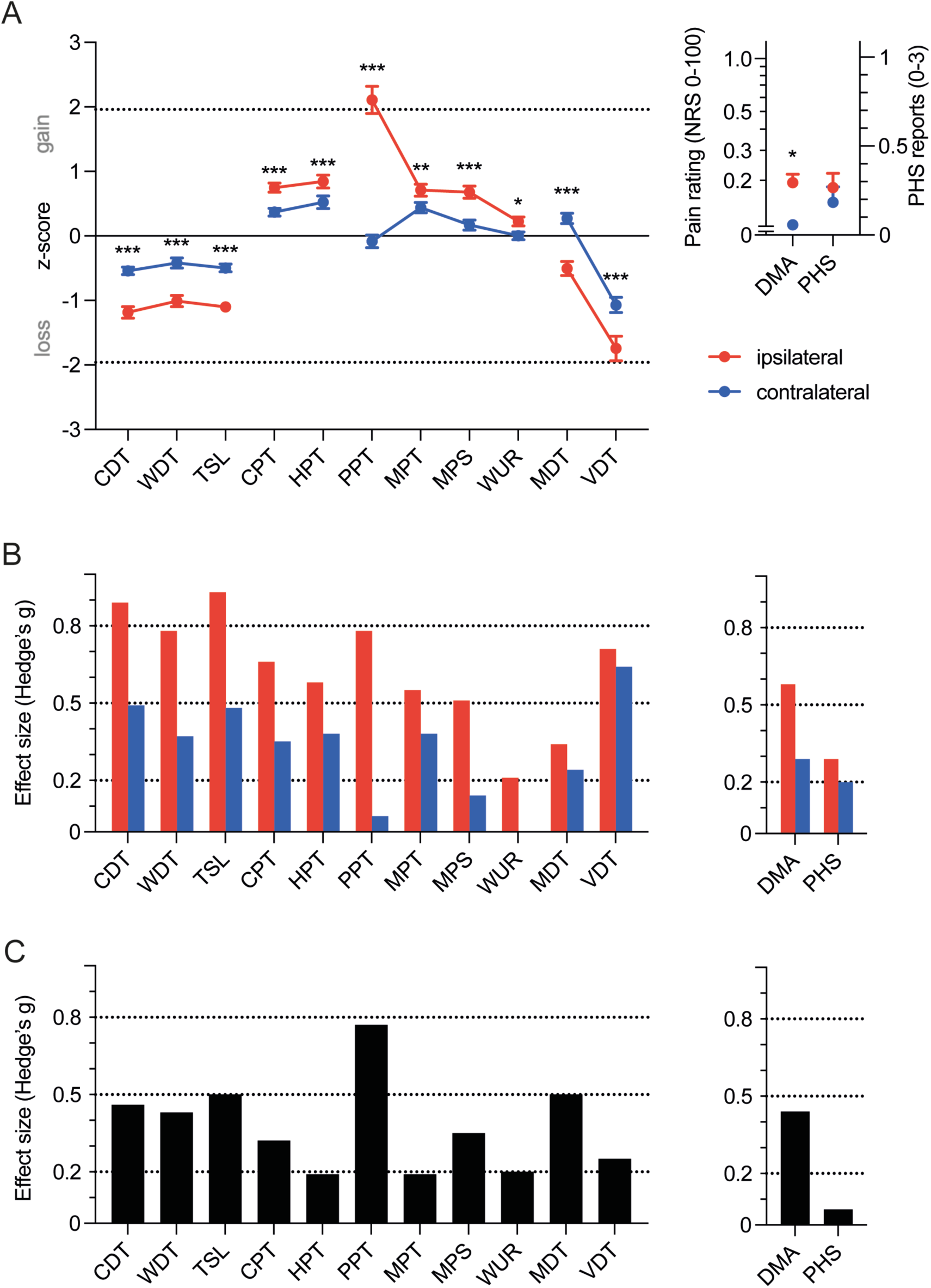
Comparison of somatosensory profiles at the affected ipsilateral and the unaffected contralateral mirror image test site. **(A)** Weighted mean z-scores for all QST parameters at the ipsilateral and contralateral test sites across all studies reporting data for both QST sites (14 studies, 1194 CRPS patients; weighted mean ± 95% confidence interval). DMA and PHS are reported as absolute values as z-transformation is not possible.^32^ Dashed lines indicate deviation of 1.96 standard deviations from the mean of the reference population. Asterisks indicate statistical significance for ipsilateral vs. contralateral comparison (*** *P* < 0.001). **(B)** Effect size (Hedges’ g) for the change relative to the healthy DFNS reference population for ipsilateral and contralateral test sites. **(C)** Effect size (Hedges’ g) for the difference between ipsilateral and contralateral QST assessments. Dashed lines indicate the distinction between no relevant difference and weak effect size (*g* = 0.2), between weak and moderate effect size (*g* = 0.5) and between moderate and strong effect size (*g* = 0.8). CDT: cold detection threshold, WDT: warm detection threshold, TSL: thermal sensory limen, CPT: cold pain threshold, HPT: heat pain threshold, PPT: pressure pain threshold, MPT: mechanical pain threshold, MPS: mechanical pain sensitivity, WUR: wind-up ratio, MDT: mechanical detection threshold, VDT: vibration detection threshold, DMA: dynamic mechanical allodynia, PHS: paradoxical heat sensations.

The *contralateral* sensory profile in these studies largely resembled changes at the ipsilateral site but at lower magnitude (Fig. 3A). However, this was not the case for PPT and DMA, which were both markedly less prevalent contralaterally (Fig. 3B). In the comparison between the ipsi- and contralateral QST profiles consequently only PPT (g = 0.77) reached a moderate-to-large effect size (Fig. 3C).

### Changes in somatosensory profiles across CRPS duration

We stratified the included studies by CRPS duration into early (< 6 months), intermediate (6-12 months) and late time window (> 12 months) as shown in Fig. 4A. Across all time windows thermal and mechanical detection thresholds revealed a loss of function at the ipsilateral site (Fig. 4B). Sensory loss in CDT, WDT and TSL occurred at similar magnitude and was also stable across time windows. Loss in MDT exhibited an inverse U-shape across time windows and similar magnitudes of loss in the early and late groups as in thermal detection. VDT displayed the largest sensory loss, which was, again stable across time windows (Figs. 4B and 5A).

**Figure 4.**
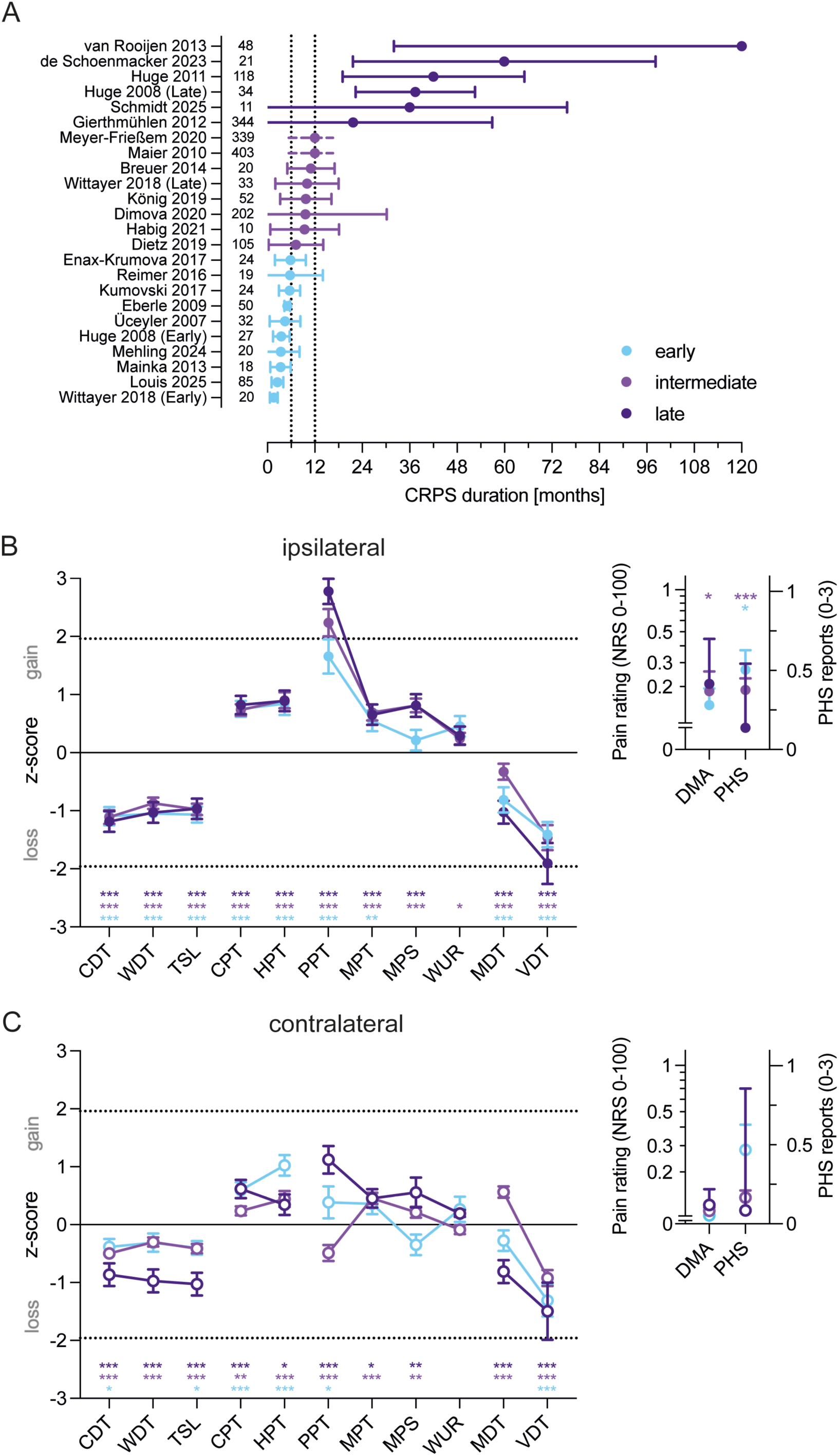
Impact of CRPS duration on somatosensory profiles. **(A)** Studies were stratified into an early (< 6 months), intermediate (6-12 months) and late (> 12 months) group by average CRPS duration of the respective studies (mean ± SD; numbers on the right indicate the numbers of participants included in each study). For Meyer-Frießem^42^ and Maier^34^ no absolute values for CRPS duration were available, so estimators based on reported numbers of patients with CRPS durations shorter or longer were used for approximation of mean duration (see methods for details). **(B)** Weighted mean z-scores of disease duration subgroups for QST in ipsilateral test sites (mean ± 95% confidence intervals). **(C)** Weighted mean z-scores of disease duration subgroups for QST in contralateral test sites (mean ± 95% confidence intervals). Dashed lines indicate deviation of 1.96 standard deviations from the mean of the reference population. * *P* < 0.05; ** *P* < 0.01; *** *P* < 0.001. CDT: cold detection threshold, WDT: warm detection threshold, TSL: thermal sensory limen, CPT: cold pain threshold, HPT: heat pain threshold, PPT: pressure pain threshold, MPT: mechanical pain threshold, MPS: mechanical pain sensitivity, WUR: wind-up ratio, MDT: mechanical detection threshold, VDT: vibration detection threshold, DMA: dynamic mechanical allodynia, PHS: paradoxical heat sensations.

**Figure 5.**
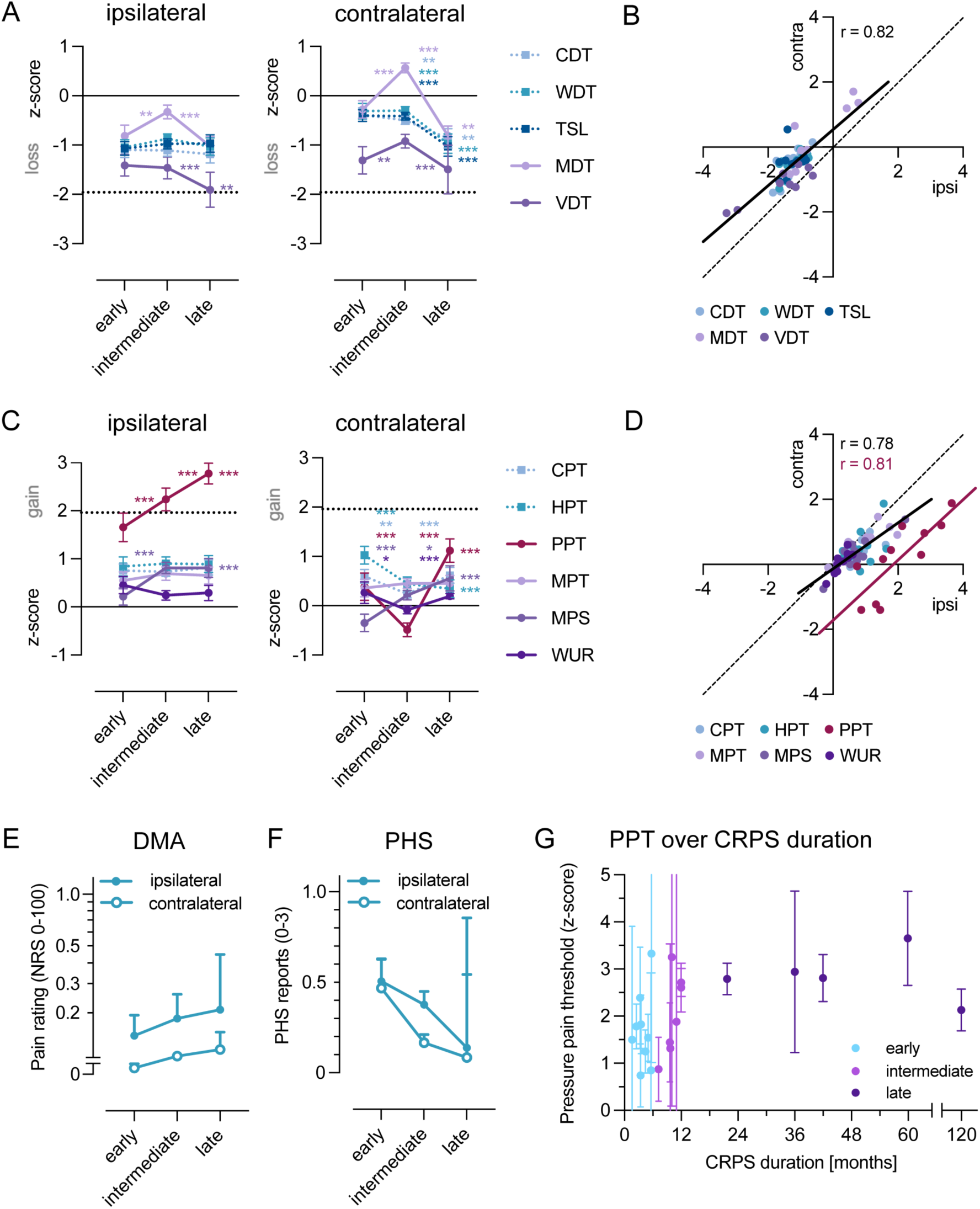
Impact of CRPS duration on individual QST parameters and the relationship between the ipsilateral (affected) site and the contralateral (mirror image) site. **(A)** Z-scores for thermal and mechanical detection parameters at the ipsi- and contralateral test sites across disease duration subgroups. **(B)** Correlation and simple linear regression of ipsi- and contralateral detection parameters. **(C)** Z-scores for thermal and mechanical nociception parameters at the ipsi- and contralateral test sites across disease duration subgroups. **(D)** Correlation and simple linear regression of ipsi- and contralateral nociceptive parameters excluding PPT (black line) and of ipsi- and contralateral PPT (red line). **(E)** Dynamic mechanical allodynia (pain to stroking light touch) across disease duration subgroups at the ipsi- and contralateral test sites. **(F)** Paradoxical heat sensations across duration subgroups at the ipsi- and contralateral test sites. Circles indicate means ± 95% confidence intervals. **(G)** Ipsilateral pressure pain threshold as reported in studies included in stratification by CRPS duration. Mean ± 95% confidence intervals. Asterisks on lines indicate statistical significance comparing the respective time windows of disease duration, i.e. early vs. intermediate, or intermediate vs. late. Asterisks on the right side of a circle indicate statistical significance comparing early and late time windows. * *P* < 0.05; ** *P* < 0.01; *** *P* < 0.001. CDT: cold detection threshold, WDT: warm detection threshold, TSL: thermal sensory limen, CPT: cold pain threshold, HPT: heat pain threshold, PPT: pressure pain threshold, MPT: mechanical pain threshold, MPS: mechanical pain sensitivity, WUR: wind-up ratio, MDT: mechanical detection threshold, VDT: vibration detection threshold, DMA: dynamic mechanical allodynia, PHS: paradoxical heat sensations.

Contralaterally, sensory loss in detection thresholds was strongest in the late group (Fig. 4C). While CDT, WDT and TSL showed no or mild sensory loss at the contralateral site in the early and intermediate group, loss in the late group increased and reached the same magnitude as in the affected area. Contralateral MDT exhibited a similar inverse U-shape across time windows as in the affected side and aggravated to the level of the affected side in the late group (Fig. 5A). Sensory loss in VDT resembled VDT in the affected area across subgroups (Fig. 5A, supplement 6). Overall, ipsilateral non-nociceptive QST parameters were shifted leftwards by approximately 0.5-1 z vs. contralateral estimates, i.e., towards more sensory loss (Fig. 5B).

As outlined, *nociceptive* parameters displayed a general gain of function across all subgroups in the affected area (Fig. 4B). Nociceptive gain only slightly increased across time brackets for CPT, HPT and MPS ipsilaterally, and remained unchanged for MPT and WUR. PPT however showed a drastic gain already in the early time window, which massively further increased in both the intermediate and the late time window (Figs. 4B and 5C).

At the contralateral site sensory gain was not prevalent in all subgroups (Fig. 4C). While HPT showed mild sensory gain that did not change across subgroups, CPT did not show sensory gain in the early group but progressive gain in the intermediate and late group. MPT displayed stable sensory gain across subgroups. MPS did not differ from the reference population in the early group but developed progressive gain in the intermediate and late group (Figs. 4B and 5C). WUR did not significantly differ from the reference population in the early and intermediate group and showed a slight gain in the late group. PPT showed a U-shaped course with statistically insignificant trend towards a slight gain in the early group, a slight loss in the intermediate group and then clear sensory gain in the late group (+1.10 ± 0.15, *P* < 0.0001; Figs. 4C and 5C; supplement 6). There was an overall close correlation between ipsi- and contralateral nociceptive QST parameters with contralateral magnitudes of about 2/3 of the ipsilateral ones (Fig. 5D). In contrast, PPT exhibited a parallel rightwards-shifted relationship with approximately 2.2 z lower z-scores contralaterally (Fig. 5D).

DMA was prevalent ipsilaterally across all subgroups but not contralaterally (Figs. 4B, 4C and 5E). PHS was present both in the early and intermediate group with a higher magnitude at the affected than unaffected site (Figs. 4B, 4C and Fig. 5F). PHS at the ipsilateral affected site decreased from the early to late group although the magnitude of sensory loss remained unchanged. Eventually, PHS in the late group became near zero and did not differ significantly from the reference population (Fig. 5F).

Comparison of the different QST assessments revealed pressure pain thresholds as the most pronounced abnormality that was highly significantly more abnormal than any other QST parameter; pressure hyperalgesia was, therefore, more pronounced than any other hyperalgesia finding. Plotting the average PPT over the average disease duration of the respective study disclosed that in early stages of the disease pressure hyperalgesia was present but almost always modest, becoming variable in a transitional time window and eventually showing massive pressure hyperalgesia in any of the late CRPS studies (Fig. 5G). Notably, for PPT, we found a strong correlation between ipsilateral and contralateral test sites (Pearson *r* = 0.81) with a parallel upward shift of the ipsilateral PPT z-scores by approximately two z-values towards more sensory gain (Fig. 5D). Accordingly, pressure hyperalgesia was lower at the contralateral site irrespective of the disease duration in the respective study, which is consistent with the finding that significant contralateral pressure hyperalgesia was encountered only in some studies in the intermediate time window but in all studies in the late time window (Fig. 5D).

### Mean ongoing pain modestly influences some somatosensory profiles

We stratified studies by mean ongoing pain ratings into < 5 and ≥ 5 on an 11-point numerical rating scale (0-10). There was no appreciable difference between groups in thermal detection as well as thermal and punctate pain. In contrast, pressure hyperalgesia in the high pain rating group was significantly higher than in the low pain rating group (2.48 ± 2.96 vs 2.04 ± 5.20 z, *P* < 0.01). Likewise, sensory loss was significantly larger in the high pain group for tactile (-0.65 ± 1.97 vs. -0.24 ± 2.80 vs. z, *P* < 0.01) and vibration detection thresholds (-1.75 ± 3.97 vs. -1.20 ± 3.83 vs. z, *P* < 0.001; Fig. 6). Thus, a similar magnitude of enhanced mechanoreceptive hypoesthesia paralleled inflated pressure hyperalgesia. However, effect sizes were below meaningful levels (Fig. 6B).

**Figure 6.**
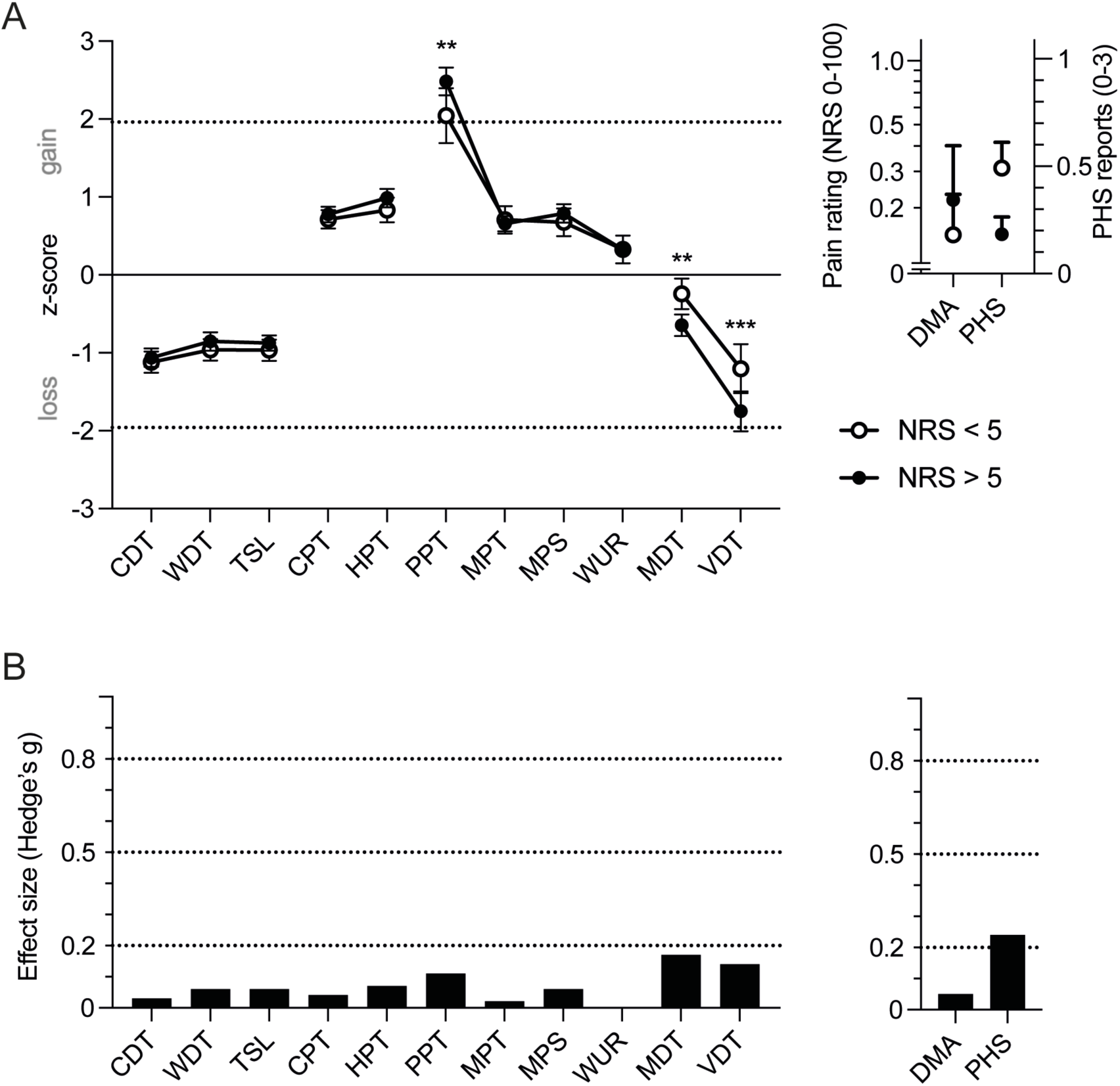
Ipsilateral somatosensory profiles of studies stratified for mean pain ratings (NRS) < 5 and ≥ 5. **(A)** Weighted mean z-scores for all QST parameters stratified by mean pain ratings on a numerical rating scale from 0-10. Weighted mean ± 95% confidence intervals. DMA and PHS are reported as absolute values as z-transformation is not possible.^32^ Dashed lines indicate deviation of 1.96 standard deviations from the mean of the reference population. **(B)** Effect size (Hedges’ g) for the difference between the NRS < 5 and NRS ≥ 5 groups. Dashed lines indicate the distinction between no relevant difference and weak effect size (*g* = 0.2), between weak and moderate effect size (*g* = 0.5), and between moderate and strong effect size (*g* = 0.8). * *P* < 0.05; ** *P* < 0.01; *** *P* < 0.001. CDT: cold detection threshold, WDT: warm detection threshold, TSL: thermal sensory limen, CPT: cold pain threshold, HPT: heat pain threshold, PPT: pressure pain threshold, MPT: mechanical pain threshold, MPS: mechanical pain sensitivity, WUR: wind-up ratio, MDT: mechanical detection threshold, VDT: vibration detection threshold, DMA: dynamic mechanical allodynia, PHS: paradoxical heat sensations.

### Effects of differential sex composition in the disease duration subgroups

Based on the sex difference in pressure hyperalgesia described previously in a large cohort^42^ we calculated the potential drift related to an enriched female subpopulation in the intermediate and late time windows (from 71.8% to 76.4% and 78.5%, see Table 1). However, sex composition explained only a minor proportion of approximately 0.1 z-values of enhanced pressure hyperalgesia in the intermediate and late subgroups as sex-related (11% and 15%, respective of the observed increase in pressure hyperalgesia).

### QST as a measure for response to therapy

Four studies examined the response to therapeutic interventions based on the DFNS QST protocol (*n* = 66 patients).^43–46^ Three out of these four studies found a complete normalisation of pressure hyperalgesia with successful treatment; this finding was consistent with the only case report which to our knowledge describes QST before and after biologics treatment in CRPS (not included in the systematic analysis, see supplement 7).^47^

## Discussion

We reviewed sensory profiles of over 2000 CRPS patients assessed in studies adhering to the standardised DFNS QST protocol. These testing protocols provide comprehensive information about somatosensory function supporting mechanism-based management stratification.^27,30,31,48^ QST is often used to assess nerve-damage-related pain conditions (neuropathic pain), whereas CRPS is *chronic primary pain*, caused by nerve *dysfunction* rather than damage^14^; over the past two decades CRPS has constituted the most extensively DFNS-QST assessed non-neuropathic pain condition.^33^ Similar to neuropathic pain, in CRPS incomplete understanding of the underlying pathophysiological abnormalities hampers therapeutic progress, and there is a lack of objective diagnostic criteria. Our quantitative analysis of QST data published over the past two decades clarifies the somatosensory profile presented by this patient group including for the first time dramatic changes associated with disease duration. Our results permit inferences about disease mechanisms and future stratification approaches towards effective treatments.

### Moderate global detection deficits

Significant blunting of all tactile and thermal detection modalities over the affected body part, i.e., the distal extremity^9^, is evident. However, the degree of blunting in CRPS is much smaller than that reported in neuropathic pain conditions, which in turn feature only minor inter-condition variability for this feature.^30,34^

### Positive signs are prominent and most marked for PPT

There is significant hyperalgesia encompassing all nociceptive modalities; these abnormalities are more pronounced than reported in neuropathic pain.^30,34^ Pressure pain threshold, which identifies deep tissue hyperalgesia, is most clearly prominent (> 2 SD above reference data), statistically amounting to more than half of all CRPS patients displaying abnormal pressure pain sensitivity. We found that hyperalgesia to blunt pressure is further distinguished from other QST parameters by its pronounced progression from early to late CRPS.

In other words, adult CRPS patients have a very high likelihood of *abnormal* pressure pain sensitivity at *any* time in their disease trajectory. Already in the early time window 26% of CRPS patients are expected to have abnormally high ipsilateral pressure pain sensitivity (i.e., exceeding the 95% CI of the reference range of healthy subjects), which then further expands to 57% and 78% in the intermediate and late time windows.

Pressure hyperalgesia in the affected limb in CRPS has traditionally been quantified by ipsilateral to contralateral comparison, as opposed to using healthy reference values.^49^ One important finding from this analysis is that such a within-patient approach of analysing QST is valid for assessment at any time point, as patients are more than 2 standard deviations more sensitive ipsilaterally than contralaterally in all studies that provided these data (Fig. 5D). At the same time, however, within-patient comparison therefore fails to identify the profound step-increase of pressure hyperalgesia between 4-12 months which we have demonstrated (Fig. 5G).

Whereas transition from modest to strong pressure hyperalgesia appears to occur relatively early in the ipsilateral limb, beginning at around four months after CRPS onset and being fully evident by the end of the first year, pressure hyperalgesia appears to then plateau and not further increase for time windows up to ten years CRPS duration. The evidence for this ‘PPT step change’ is formally limited by the cross-sectional nature of the available data. Nevertheless, the observed peculiar, unexpected, and nearly isolated pressure pain ‘progression’ pattern is supported by analysis of all studies.

### Abnormal sensitivity in additional body areas is dynamic over time

*Sensory loss* of both thermal and mechanical detection on the *contralateral* side, which is very modest during the first year after CRPS onset enhances in later CRPS, to almost the same magnitude as in the ipsilateral affected area (Fig. 5A), an effect already reported in early studies.^38,50^ Whereas contralateral loss of detection has also been reported in neuropathic pain conditions, for example after traumatic nerve injury, time course data are not available for these neuropathy cohorts.^44,51^

In later CRPS (> 12 months), *pressure hyperalgesia* in the non-painful, *contralateral* mirror image test site becomes highly significant versus healthy normal. Mirror-image findings in our analysis are consistent with the pivotal early (non-DFNS) study by Veldman^3^. Our results suggest that pressure hyperalgesia in later CRPS may even further extend to more remote test sites, suggesting prevalence of widespread-rather than only mirror-contralateral sensitivity spread patterns; however this information is based on data from only two studies.^41,52^ These pattern of sensitivity extensions (mirror or beyond) are incongruent with the classical pattern of central sensitisation (secondary hyperalgesia), which would remain unilateral and confined to close-by segments.^53^

The observed expansion of the territory of pressure hyperalgesia in late CRPS beyond the initially affected area represents a partial loss of the regional disease features which define CRPS. We found that to a lesser degree such expansion also occurs in other QST modalities, both nociceptive and non-nociceptive (detection). Remote pressure hyperalgesia and tactile allodynia in CRPS may be dynamically maintained and modulated by changes of both ipsilateral stimulus-evoked pain, and nociceptive activity.^54–60^ Notably, given the prominence of PPT indicating deep tissue sensitisation, it might be speculated that in CRPS deep tissue hyperalgesia drives these changes across all modalities, mediated by crosstalk into processing of superficial cutaneous sensory input.^61^ Whether hypersensitivity expansion in individual patients heralds risk for pain spread, which is observed in later CRPS, requires further study.^62^

### Increasing sex disbalance over time

We observed that beyond the already high preponderance of female sex in CRPS there was a highly significant alteration in the contribution of both sexes over time which, to our knowledge has not been reported previously. Increasing disease duration progressively shifted the sex balance even more towards female, pointing toward differences between cohorts defining the early, intermediate or late CRPS QST profiles, respectively. In the included studies, this alteration however explained only a small proportion of the observed profound deepening of PPT over time. Longitudinal studies over at least two years, currently missing, would be required to allow clarification whether these observations indicate that spontaneous improvement rates differ between males and females (female sex being a risk factor), or alternatively, whether the observed shift in sex proportions is due to recruitment-related factors.

### Implications for disease mechanisms, disease classification, and rehabilitative treatment

QST profiles characterised by pressure pain, similar to those characterised by the cutaneous parameters cold and heat, are thought to point towards involvement of primary afferent-related C-fibre mechanisms, rather than central sensitisation^63–65^; our findings indicating exaggerated (deep) pressure hyperalgesia, relative to cutaneous hyperalgesia *of any modality* further suggest that CRPS differs from most neuropathic pain conditions by disproportionate, primary-afferent related sensitisation of *deep* tissue.

This particularly strong PPT abnormality might suggest the relevance of persistent pathogenic factors that affect deep tissue primary afferents, in addition to potential global dysregulation of central pain processing (chronification), for explaining these sensory profiles. The nature of these factors requires further research.

The outlined distinction of CRPS from neuropathic pain across positive signs is evident without differences between CRPS I and CRPS II^33^, a finding consistent with the recent re-classification of CRPS, including CRPS II as a chronic primary pain condition rather than neuropathic pain.^14^

CRPS has a detrimental impact on limb movement rendering rehabilitation particularly challenging.^5,66^ Although in the included studies deep pressure hyperalgesia was assessed by pressure on muscles, an effect on motor performance either directly or through concomitant irritation of fascia is unavoidable^50^; in deep muscle pressure hyperalgesia fascia are likely to be hyperalgesic and contribute to painful joint movement, a prominent and clinically problematic feature in CRPS.^67–69^ Independently, specific testing in both experimental deep tissue hyperalgesia models and in CRPS patients has suggested that (non-muscular, deep) joint tissues exhibit a strong degree of hyperalgesia or are even more hyperalgesic than muscle.^67–69^ Joint hyperalgesia was only tested directly in one of the included studies, in which it exceeded the magnitude of muscle hyperalgesia.^70^ The PPT outcomes in this study, therefore also provide new data underpinning the pragmatically observed challenges in rehabilitation settings for persistent CRPS: it is already recognised that improvement of autonomic changes in *later* CRPS such as reduced swelling does not equal pain improvement and easier rehabilitation.^5,14^ Our data underline that patients and their treatment teams will additionally often have to content with dramatically sensitised deep tissues.

### Dysesthesias

In contrast to hyperalgesias, *dysesthesias*, namely dynamic mechanical allodynia and paradoxical heat sensations, are present at a low level with a high variability in CRPS cohorts from all duration subgroups, suggesting that these signs are only present in a minority of patients; they might, of course, be substantial in this subgroup. In our clinical experience, extreme pressure hyperalgesia can be mistaken as abnormal DMA; patients frequently report that ‘touch’ is painful, but closer examination then shows that the abnormality lies with even the slightest pressure rather than touch. For neuropathic pain, it has been suggested that paradoxical heat sensations are a sequel of loss of thermal detection.^71^ However, this proposition is at variance with the CRPS data in this study, where paradoxical heat sensations reduce gradually in acute-to-late transition, while loss of thermal detection remains entirely unchanged. Further, PHS does not flare up contralaterally in late CRPS, when loss of contralateral thermal detection increases. We speculate that these observations reflect compensatory, readjusting central mechanisms, reducing the likelihood of thermal dysesthesia with progressing disease duration of CRPS. As mentioned above, change in cohort composition might be relevant.

### Prognostic relevance of the QST abnormalities

The prognosis of early CRPS of < 6-12 months duration is overall good, with four in five patients eventually being able to enjoy substantial natural improvement to a degree that they either recover entirely or experience relevant improvements in their quality of life. Spontaneous recovery thereafter becomes less likely, and is very rare > 15 months.^23^ This suggests that cross-sectional cohorts with > 6 months duration are successively enriched with patients that will not recover, and our data suggest for the first time that these patients (more often female) with little potential for recovery feature extreme pressure hyperalgesia. These results imply that an extreme PPT threshold is either an unfavourable prognostic marker already early in CRPS, or that a subgroup of patients, indistinguishable by early QST, later ‘switches’ psychophysical properties. Similarly *contralateral* mechanical punctate and pressure hyperalgesia as well as sensory loss, which statistically emerge in late CRPS might either constitute a *de novo* protracted phenotype in these patients or instead characterize a more severe phenotype already in early CRPS. Notably, their contralateral appearance seems to be linked to a higher intensity of ongoing pain.^22,72,73^

Extreme PPT *gain* has been suggested as a *poor prognostic sign* leading to a more unfavourable pain-disease trajectory in a subgroup of patients previously also identified in another pain patient cohort.^74^ Accordingly, for clarification of CRPS disease trajectory longitudinal testing is required.

Our data do not provide information about changes in spontaneous pain over time in patients who do *not* improve, although we found that patients with higher pain intensities tended to have somewhat more severe pressure hyperalgesia. The question of longitudinal changes in spontaneous pain and any relation to PPT also remains to be examined.

### Underpinning mechanisms for the PPT step

It is tempting to speculate about both the molecular mechanisms responsible for rendering PPT the ‘CRPS signature abnormality’, and those mechanisms (if different) underpinning the observed PPT step increase between about 4-12 months (Fig. 5G). Patients with early CRPS harbour pro-nociceptive IgM serum autoantibodies^75^, and the time of the PPT upward step broadly coincides with a potential immunological switch to pro-nociceptive IgG autoantibodies.^76^ However, longitudinal studies are not available. Experimental studies would be required comparing the hyperalgesia-inducing capacity of these antibodies to explore their relevance for the QST step change.

### CRPS subtyping

Distinct subtypes (or phenotypes) of CRPS have been explored e.g., warm/cold, dystonic/non-dystonic, presence/absence of contralateral abnormalities, impact of the inciting event, early/persistent and others.^18,77,78^ The validity of these proposed subtypes and how they relate to underlying pathophysiological mechanisms is unknown, however, if confirmed their understanding may inform differential treatment.^22,72,73,78,79^ There is an urgent but unmet need for better patient stratification on the background of only little solid evidence regarding both optimal pharmacological and rehabilitative treatment.^11,80^

Our data provide evidence to inform these continuing discussions^14^; they suggest that both disease duration and PPT magnitude may be assessed in longitudinal studies for their usefulness in identifying a non-resolving subtype. For clarity, such a subtype might then not be the sole consequence of late ‘chronification’ but might rather indicate specific (patho-) biological factors, which could be already present early. Should prognostic properties of excessive pressure hyperalgesia be confirmed, then this assessment parameter may aid patient information in the future. Given the potential distinct immune profile of persistent CRPS outlined above, assessment of PPT may also support enrolment into certain clinical trials and future clinical decision making.

These data also lend weight to the proposed introduction of a third CRPS subtype, which is currently being considered by the WHO.^14^ This proposal was based on the observation that autonomic signs are often so far diminished over time as to not allow the diagnosis of CRPS according to the Budapest criteria^13^ any longer, in the face of ongoing severe pain in patients who initially clearly fulfilled these criteria. Our data indicate for the first time that, counter-intuitively, patients with this proposed third subtype (which should overlap with the ‘persistent subtype’) are also likely to suffer from markedly increased pressure hyperalgesia and consequent movement restriction with challenges to rehabilitation.

Finally, our data highlight potential challenges with the validity of PPT obtained in patients with persistent CRPS; the magnitude of the PPT shift in some patients should make it near impossible to use standardised pressure algesiometry (PPT) which relies on reliably detecting a minimal change of 50 kPa. Only two of the reviewed studies reported loss of data due to QST intolerance (3/21 patients^52^ and 7/202 patients^18^), although in our experience this is a common challenge in this patient group at tertiary centres.^81^

### Implications for future longitudinal studies

Given the relatively small proportion of those patients with persistent CRPS, the feasibility of longitudinal studies starting from CRPS onset is uncertain. Prospective studies might be most successful if they focus on populations, which are at high risk for the development of CRPS, e.g., after high impact trauma and/or surgery.^6,7^ Pressure pain intensity should be included amongst predefined prognostic outcome parameter and should ideally be measured in regular intervals to enable detection of any dramatic changes.^22^

### Limitations

Lack of consistent reporting was evident across the reviewed studies hampering easy aggregation of data for quantitative analysis. Accordingly, full reporting of data and respective statistical measures of uncertainty and precision (e.g., standard deviation, confidence intervals, sensitivity analyses, effect sizes) and adequate stratification in reporting (e.g., for sex, disease duration, upper/lower extremity, etc.) is of utmost importance and consensus on criteria for optimising have been suggested recently.^82^ As not all studies reported full QST profiles, not all parameters across the profiles reported here stem from the same cohorts. Due to limited data availability, we did not stratify the data on the patient level but only on the level of included study cohorts. Thus, the reported subgroups in this study do not reflect the disease duration of all but the majority of patients included in them. Tentative recalculation of non-mean/SD data was done when only median and (interquartile) range were reported (see supplement 3). Standardised reporting would highly facilitate the use of DFNS QST data in future quantitative analyses. Further, the patient population analysed here stems mostly from tertiary care centres in Central Europe, mainly Germany. Recruitment of mild or improved cases might significantly trail recruitment of more severe cases, especially in long-standing CRPS. We do not know whether our findings are applicable to non-European populations, and Asian studies have shown a higher proportion of lower limb features and even suggested modification of diagnostic criteria.^83–85^

### Summary

The results of this systematic review and evidence synthesis of DFNS-QST-studied sensory profiles in CRPS (I and II) patients indicate clearly distinct patterns from neuropathic pain conditions, supporting the recent reclassification of all CRPS as chronic primary pain. Pressure pain threshold (PPT) is the CRPS signature abnormality, which demonstrates a unique and dramatic step change between 4-12 months disease duration. Both PPT and the CRPS disease duration may be suited as prognostic markers and CRPS subgrouping parameters.

## Data Availability

All data produced in the present study are available upon reasonable request to the authors.

