## Supplementary Materials for "Phenotype Progression of Complex Regional Pain Syndrome Identified by Quantitative Sensory Testing"

**Supplement 1:**

**Test stimuli and somatosensory modalities used in QST assessment following the DFNS protocol**

| Type of Stimulus | QST Modality | Devices used for QST Testing | Peripheral Nerve fibre tested | Central Pathway |
|---|---|---|---|---|
| Thermal | Cold Detection Threshold (CDT) | Contact thermal testing devices (Computer-controlled ramp rates) | Aδ cold | Spinothalamic |
| | Warm Detection Threshold (WDT) | | C warm | Spinothalamic |
| | Cold Pain Threshold (CPT) | | C | Spinothalamic |
| | Heat Pain Threshold (HPT) | | C | Spinothalamic |
| | Thermal Sensory Limen (TSL) | | Aδ cold, C warm | Spinothalamic |
| | Paradoxical Heat Sensation (PHS) | | undefined | Spinothalamic |
| Mechanical Touch | Mechanical Detection Threshold (MDT) | Calibrated von Frey Filaments | Aβ-LTM | Lemniscal |
| Mechanical Pain | Mechanical Pain Threshold (MPT) | Calibrated Pinprick needles | Aδ-HTM | Spinothalamic |
| | Mechanical Pain Sensitivity (MPS) | | Aδ-HTM | Spinothalamic |
| | Dynamic Mechanical Allodynia (DMA) | Cotton wool, Q-tip and brush | Aβ-LTM | Spinothalamic |
| | Wind up (WUR) | Calibrated pinprick needles | Aδ, C | Spinothalamic |
| Vibration | Vibration Detection Threshold (VDT) | 64 Hz Tuning fork | Aβ-LTM | Lemniscal |
| Blunt Pressure | Pressure Pain Threshold (PPT) | Pressure algometer | Aδ, C deep | Spinothalamic |

Stimuli, corresponding QST parameters and corresponding peripheral nerve fibres and central nervous system pathways. Thick or thin myelinated fibres (Aβ, Aδ) involved in light touch, vibration, thermal sensations (cold) and first pain; fine unmyelinated fibres (C) involved in temperature sensation (warm) and second pain. LTM or HTM low threshold or high threshold mechanoreceptors.

**Supplement 2:**

**Publications with QST assessment according to the DFNS protocol and included in quantitative analysis (list of QST parameters reported in the respective publication) (n=23)**

Breuer AJ, Mainka T, Hansel N, Maier C, Krumova EK. Short-term treatment with parecoxib for complex regional pain syndrome: a randomized, placebo-controlled double-blind trial.Pain Physician. 2014;17:127-37. (reported HPT, PPT)

De Schoenmacker I, Mollo A, Scheuren PS, Sirucek L, Brunner F, Schweinhardt P, Curt A, Rosner J, Hubli M. Central sensitization in CRPS patients with widespread pain: A cross-sectional study. Pain Med 2023;24:974-984. (reported full QST profile)

Dietz C, Müller M, Reinhold AK, Karch L, Schwab B, Forer L, Vlckova E, Brede EM, Jakubietz R, Üçeyler N, Meffert R, Bednarik J, Kress M, Sommer C, Dimova V, Birklein F, Rittner HL. What is normal trauma healing and what is complex regional pain syndrome I? An analysis of clinical and experimental biomarkers. Pain 2019;160:2278-2289. (reported CDT, WDT, TSL, CPT, HPT, PPT, MPT, MPS, WUR, MDT, VDT)

Dimova V, Herrnberger MS, Escolano-Lozano F, Rittner HL, Vlckova E, Sommer C, Maihöfner C, Birklein F. Clinical phenotypes and classification algorithm for complex regional pain syndrome. Neurology 2020;94:e357-e367. (reported full QST profile)

Eberle T, Doganci B, Krämer HH, Geber C, Fechir M, Magerl W, Birklein F. Warm and cold complex regional pain syndromes: differences beyond skin temperature? Neurology 2009;72:505-512 (reported full QST profile)

Enax-Krumova EK, Lenz M, Frettlöh J, Höffken O, Reinersmann A, Schwarzer A, Westermann A, Tegenthoff M, Maier C. Changes of the Sensory Abnormalities and Cortical Excitability in Patients with Complex Regional Pain Syndrome of the Upper Extremity After 6 Months of Multimodal Treatment. Pain Med 2017;18:95-106 (reported CDT, WDT, TSL, CPT, HPT, PPT, MPT, MPS, WUR, MDT, VDT)

Gierthmühlen J, Maier C, Baron R, Tölle T, Treede RD, Birbaumer N, Huge V, Koroschetz J, Krumova EK, Lauchart M, Maihöfner C, Richter H, Westermann A and German Research Network on Neuropathic Pain (DFNS) study group. Sensory signs in complex regional pain syndrome and peripheral nerve injury. Pain 2012;153:765-774 (reported full QST profile)

Habig, K., Lautenschlager, G., Maxeiner, H., Birklein, F., Krämer, H.H., and Seddigh, S. Low mechano-afferent fibers reduce thermal pain but not pain intensity in CRPS. BMC Neurol 2021;21:272 (reported CDT, WDT, TSL, CPT, HPT, MPT, MPS)

Huge V, Lauchart M, Förderreuther S, Kaufhold W, Valet M, Azad SC, Beyer A, Magerl W. Interaction of hyperalgesia and sensory loss in complex regional pain syndrome type I (CRPS I). PLoS One 2008;3:e2742 (reported CDT, WDT, TSL, CPT, HPT, PHS)

Huge V, Lauchart M, Magerl W, Beyer A, Moehnle P, Kaufhold W, Schelling G, Azad SC. Complex interaction of sensory and motor signs and symptoms in chronic CRPS. PLoS One 2011;6:e18775 (reported full QST profile)

König S, Bayer M, Dimova V, Herrnberger M, Escolano-Lozano F, Bednarik J, Vlckova E, Rittner H, Schlereth T, Birklein F. The serum protease network-one key to understand complex regional pain syndrome pathophysiology. Pain 2019;160:1402-1409 (reported CDT, WDT, TSL, CPT, HPT, PPT, MPT, MPS, WUR, MDT, VDT, PHS)

Kumowski N, Hegelmaier T, Kolbenschlag J, Maier C, Mainka T, Vollert J, Enax-Krumova E. Unimpaired endogenous pain inhibition in the early phase of complex regional pain syndrome. Eur J Pain 2017;21:855-865 (reported full QST profile)

Louis MH, Legrain V, Aron V, Filbrich L, Henrard S, Barbier O, Libouton X, Mouraux D, Lambert J, Berquin A. Early CRPS Is a Heterogeneous Condition: Results From a Latent Class Analysis. Eur J Pain. 2025;29:e4785 (reported full QST profile)

Maier C, Baron R, Tölle TR, Binder A, Birbaumer N, Birklein F, Gierthmühlen J, Flor H, Geber C, Huge V, Krumova EK, Landwehrmeyer GB, Magerl W, Maihöfner C, Richter H, Rolke R, Scherens A, Schwarz A, Sommer C, Tronnier V, Üçeyler N, Valet M, Wasner G, Treede DR. Quantitative sensory testing in the German Research Network on Neuropathic Pain (DFNS): somatosensory abnormalities in 1236 patients with different neuropathic pain syndromes. Pain 2010;150:439-450 (reported full QST profile)

Mainka T, Bischoff FS, Baron R, Krumova EK, Nicolas V, Pennekamp W, Treede RD, Vollert J, Westermann A, Maier C. Comparison of muscle and joint pressure-pain thresholds in patients with complex regional pain syndrome and upper limb pain of other origin. Pain 2014;155:591-597 (reported PPT)

Mehling K, Becker J, Chen J, Scriba S, Kindl G, Jakubietz R, Sommer C, Hartmannsberger B, Rittner HL. Bilateral deficiency of Meissner corpuscles and papillary microvessels in patients with acute complex regional pain syndrome. Pain. 2024;165:1613-1624 (reported CDT, WDT, TSL, CPT, HPT, PPT, MPT, MPS, WUR, MDT, VDT)

Meyer-Frießem CH, Attal N, Baron R, Bouhassira D, Finnerup NB, Freynhagen R, Gierthmühlen J, Haanpää M, Hansson P, Jensen TS, Kemp H, Kennedy D, Leffler AS, Rice ASC, Segerdahl M, Serra J, Sindrup S, Solà R, Tölle T, Schuh-Hofer S, Treede RD, Pogatzki-Zahn E, Maier C, Vollert J. Pain thresholds and intensities of CRPS type I and neuropathic pain in respect to sex. Eur J Pain 2020;24:1058-1071 (reported full QST profile)

Reimer M, Rempe T, Diedrichs C, Baron R, Gierthmühlen J. Sensitization of the Nociceptive System in Complex Regional Pain Syndrome. PLoS One 2016;11:e0154553 (reported full QST profile)

Schmidt H, Drusko A, Renz MP, et al. Application of the grading system for “nociplastic pain” in chronic primary and chronic secondary pain conditions: a field study. Pain 2025;166:196 (reported CDT, WDT, TSL, HPT, PPT, MPT, WUR, MDT; MPS was excluded as testing did not adhere to the DFNS protocol for MPS)

Uceyler N, Eberle T, Rolke R, Birklein F, Sommer C. Differential expression patterns of cytokines in complex regional pain syndrome. Pain 2007;132:195-205 (reported full QST profile)

van Rooijen DE, Marinus J, Schouten AC, Noldus LP, van Hilten JJ. Muscle hyperalgesia correlates with motor function in complex regional pain syndrome type 1. J Pain 2013;14:446-454 (CRPS affected site; reported CDT, WDT, CPT, HPT, PPT, WUR, VDT)

van Rooijen DE, Marinus J, van Hilten JJ. Muscle hyperalgesia is widespread in patients with complex regional pain syndrome. Pain. 2013;154:2745-2749 (CRPS contralateral and remote site; reported CDT, WDT, CPT, HPT, PPT, VDT).

Wittayer M, Dimova V, Birklein F, Schlereth T. Correlates and importance of neglect-like symptoms in complex regional pain syndrome. Pain 2018;159:978-986 (reported CDT, WDT, TSL, CPT, HPT, PPT, MPT, MPS, MDT, VDT)

**Publications with QST assessment adhering to the DFNS protocol but NOT included in quantitative analysis, with justification (n=10)**

Allmendinger F, Scheuren PS, De Schoenmacker I, Brunner F, Rosner J, Curt A, Hubli M. Contact-Heat Evoked Potentials: Insights into Pain Processing in CRPS Type I. J Pain Res. 2024;17:989-1003 (limited DFNS type QST protocol – only HPT, MPT, MPS - in a patient population, for which full QST profile data have been reported before, cf. de Schoenmacker 2023).

Bernateck M, Rolke R, Birklein F, Treede RD, Fink M, Karst M. Successful intravenous regional block with low-dose tumor necrosis factor-alpha antibody infliximab for treatment of complex regional pain syndrome 1. *Anesth Analg*. 2007;105:1148-51 (complete DFNS type QST protocol, but only a case report).

Birklein F, Riedl B, Sieweke N, Weber M, Neundörfer B. Neurological findings in complex regional pain syndromes--analysis of 145 cases. Acta Neurol Scand 2000;101:262-269 (limited DFNS type QST protocol - CDT, WDT, CPT, HPT - in a pre-DFNS cohort and assessment conformable with DFNS standard assessment, but reporting prevented transformation into standard values).

De Schoenmacker I, Sirucek L, Scheuren PS, Lütolf R, Gorrell LM, Brunner F, Curt A, Rosner J, Schweinhardt P, Hubli M. Sensory phenotypes in complex regional pain syndrome and chronic low back pain-indication of common underlying pathomechanisms. Pain Rep. 2023;8:e1110 (complete DFNS type QST protocol but reporting only condensed “meta-parameters” not allowing to extract individual QST parameters).

Dietz C, Reinhold AK, Escolano-Lozano F, Mehling K, Forer L, Kress M, Üçeyler N, Sommer C, Dimova V, Birklein F, Rittner HL. Complex regional pain syndrome: role of contralateral sensitisation. Br J Anaesth 2021;127:e1-e3 (limited DFNS type QST protocol – only CDT, WDT, CPT, HPT, PPT, MPT, MDT – in a patient population, for which QST data have been reported before, cf. Dietz 2019).

Escolano-Lozano F, Gries E, Schlereth T, Dimova V, Baka P, Vlckova E, König S, Birklein F. Local and Systemic Expression Pattern of MMP-2 and MMP-9 in Complex Regional Pain Syndrome. J Pain 2021;22:1294-1302 (complete DFNS type QST protocol was assessed but not reported in the publication).

Kinfe T, von Willebrand N, Stadlbauer A, Buchfelder M, Yearwood TL, Muhammad S, Chaudhry SR, Gravius S, Randau T, Winder K, Maihöfner C, Gravius N, Magerl W. Quantitative sensory phenotyping in chronic neuropathic pain patients treated with unilateral L4-dorsal root ganglion stimulation. J Transl Med. 2020;18:403 (complete DFNS type QST protocol in test sites at the knee, an uncommon and more proximal test area, for which no reference data were available, and thus no standard values have been reported).

Lunden LK, Kleggetveit IP, Schmelz M, Jorum E. Cold allodynia is correlated to paroxysmal and evoked mechanical pain in complex regional pain syndrome (CRPS). Scand J Pain. 2022;22:533-542 (limited DFNS type QST protocol employing the same stimulation parameters - only CDT, WDT, CPT, HPT - using a test device and assessment parameters that were conformable with DFNS reference and standard assessment, but reporting prevented transformation into standard values).

Reinhold AK, Kindl GK, Dietz C, Scheu N, Mehling K, Brack A, Birklein F, Rittner HL. Molecular and clinical markers of pain relief in complex regional pain syndrome: An observational study. Eur J Pain 2023;27:278-288 (complete DFNS type QST protocol in a patients population, for which full QST profile data have been reported before, cf. Dietz 2019).

Scheuren PS, De Schoenmacker I, Rosner J, Brunner F, Curt A, Hubli M. Pain-autonomic measures reveal nociceptive sensitization in complex regional pain syndrome. Eur J Pain 2023;27:72-85 (complete DFNS type QST protocol, but sparse reporting not allowing normalization to standard values).

**Publications with QST assessment but NOT adhering to the DFNS protocol and therefore NOT included in meta-analysis (n=15)**

Adami G, Fassio A, Rossini M, Montanari F, Manfrè S, Bonasera G, Bertelle D, Benini C, Viapiana O, Braga V, Gatti D. Long-term effectiveness and predictors of bisphosphonate treatment in type I complex regional pain syndrome. Clin Exp Rheumatol 2024;4:961-966.

Drummond PD, Finch PM. A disturbance in sensory processing on the affected side of the body increases limb pain in complex regional pain syndrome. Clin J Pain. 2014;30:301-6.

Drummond PD, Finch PM, Birklein F, Stanton-Hicks M, Knudsen LF. Hemisensory disturbances in patients with complex regional pain syndrome. Pain. 2018;159:1824-32.

Edinger L, Schwartzman RJ, Ahmad A, Erwin K, Alexander GM. Objective sensory evaluation of the spread of complex regional pain syndrome. Pain Physician. 2013;16:581-91.

Eisenberg E, Backonja MM, Fillingim RB, Pud D, Hord DE, King GW, Stojanovic MP. Quantitative sensory testing for spinal cord stimulation in patients with chronic neuropathic pain. Pain Pract. 2006;6:161-5.

Grothusen JR, Alexander G, Erwin K, Schwartzman R. Thermal pain in complex regional pain syndrome type I. Pain Physician. 2014;17:71-9.

Hartrick CT, Kovan JP, Naismith P. Outcome prediction following sympathetic block for complex regional pain syndrome. Pain Pract. 2004;4:222-8.

Kemler MA, Reulen JP, van Kleef M, Barendse GA, van den Wildenberg FA, Spaans F. Thermal thresholds in complex regional pain syndrome type I: sensitivity and repeatability of the methods of limits and levels. Clin Neurophysiol. 2000;111:1561-8.

Kemler MA, Schouten HJ, Gracely RH. Diagnosing sensory abnormalities with either normal values or values from contralateral skin: comparison of two approaches in complex regional pain syndrome I. Anesthesiology. 2000;93:718-27.

Kriek N, de Vos CC, Groeneweg JG, Baart SJ, Huygen F. Allodynia, Hyperalgesia, (Quantitative) Sensory Testing and Conditioned Pain Modulation in Patients With Complex Regional Pain Syndrome Before and After Spinal Cord Stimulation Therapy. Neuromodulation. 2023;26:78-86

Munts AG, van Rijn MA, Geraedts EJ, van Hilten JJ, van Dijk JG, Marinus J. Thermal hypesthesia in patients with complex regional pain syndrome related dystonia. J Neural Transm (Vienna). 2011;118:599-603.

Palmer S, Bailey J, Brown C, Jones A, McCabe CS. Sensory Function and Pain Experience in Arthritis, Complex Regional Pain Syndrome, Fibromyalgia Syndrome, and Pain-Free Volunteers: A Cross-Sectional Study. Clin J Pain. 2019;35:894-900.

Rommel O, Malin JP, Zenz M, Jänig W. Quantitative sensory testing, neurophysiological and psychological examination in patients with complex regional pain syndrome and hemisensory deficits. Pain. 2001;93:279-93.

Terkelsen AJ, Gierthmühlen J, Finnerup NB, Hojlund AP, Jensen TS. Bilateral hypersensitivity to capsaicin, thermal, and mechanical stimuli in unilateral complex regional pain syndrome. Anesthesiology. 2014;120:1225-36.

Vaneker M, Wilder-Smith OH, Schrombges P, de Man-Hermsen I, Oerlemans HM. Patients initially diagnosed as 'warm' or 'cold' CRPS 1 show differences in central sensory processing some eight years after diagnosis: a quantitative sensory testing study. Pain. 2005;115:204-11.

**<u>Supplement 3:</u> Calculation of mean and SD of QST parameters from median and range (or interquartile range) of QST parameters and duration of disease**

Some of the QST data used in the quantitative analysis have only been reported as median and range (Breuer et al. 2014, Mehling et al. 2024) or median and interquartile range (van Rooijen et al. 2013). Some studies reported disease duration as median range (Breuer et al. 2014, Enax-Krumova et al. 2017, de Schoenmacker et al. 2023) or median and interquartile range (Wittayer et al. 2018). For these data sets, parametric means and SDs have been derived by standard transforming equation (see below).

For data reported as median and range, mean and standard deviation were estimated according to Hozo and colleagues (Hozo et al. 2005). The approximated mean was estimated as follows with a being the minimum, b being the maximum, m being the median and n being the sample size:

$$\bar{x} = \frac{a + 2m + b}{4} + \frac{a - 2m + b}{4n}$$

The approximated standard deviation was estimated as follows:

$$SD = \frac{b - a}{4}$$

For data reported as median and interquartile range, mean and standard deviation were estimated according to Wan and colleagues (Wan et al.2014). The mean was estimated as follows with $q_1$ being the lower quartile and $q_3$ being the upper quartile:

$$\bar{x} = \frac{q_1 + m + q_3}{3}$$

The standard deviation was estimated as follows with $\eta(n)$ as provided by Wan and colleagues:

$$S = \frac{q_3 - q_1}{\eta(n)}$$

Two publications (Maier et al. 2010, Meyer-Frießem et al. 2019) did not provide absolute descriptors of CRPS duration but only the number of patients included with a duration below or above one year were reported, namely n=214 < 1 year and n=189 > 1 year (Maier et al. 2010) and n=179< 1 year, n=157 > 1 year and n=3 with unknown disease duration (Meyer-Frießem et al. 2019). Since more patients in both studies exhibited disease duration shorter than one year i.e., the median was < 1 year we decided to stratify them as belonging to the intermediate stratum.

<u>References:</u>

Breuer AJ, Mainka T, Hansel N, Maier C, Krumova EK. Short-term treatment with parecoxib for complex regional pain syndrome: a randomized, placebo-controlled double-blind trial. Pain Physician 2014;17:127-137.

De Schoenmacker I, Mollo A, Scheuren PS, Sirucek L, Brunner F, Schweinhardt P, Curt A, Rosner J, Hubli M. Central sensitization in CRPS patients with widespread pain: A cross-sectional study. Pain Med 2023;24:974-984.

Enax-Krumova EK, Lenz M, Frettlöh J, Höffken O, Reinersmann A, Schwarzer A, Westermann A, Tegenthoff M, Maier C. Changes of the Sensory Abnormalities and Cortical Excitability in Patients with Complex Regional Pain Syndrome of the Upper Extremity After 6 Months of Multimodal Treatment. Pain Med 2017;18:95-106.

Hozo SP, Djulbegovic B, Hozo I. Estimating the mean and variance from the median, range, and the size of a sample. BMC Medical Research Methodology 2005;5:13.

Maier C, Baron R, Tölle TR, Binder A, Birbaumer N, Birklein F, Gierthmühlen J, Flor H, Geber C, Huge V, Krumova EK, Landwehrmeyer GB, Magerl W, Maihöfner C, Richter H, Rolke R, Scherens A, Schwarz A, Sommer C, Tronnier V, Üçeyler N, Valet M, Wasner G, Treede DR. Quantitative sensory testing in the German Research Network on Neuropathic Pain (DFNS): somatosensory abnormalities in 1236 patients with different neuropathic pain syndromes. Pain 2010;150:439-450

Mehling K, Becker J, Chen J, Scriba S, Kindl G, Jakubietz R, Sommer C, Hartmannsberger B, Rittner HL. Bilateral deficiency of Meissner corpuscles and papillary microvessels in patients with acute complex regional pain syndrome. Pain 2024;165:1613-1624.

Meyer-Frießem CH, Attal N, Baron R, Bouhassira D, Finnerup NB, Freynhagen R, Gierthmühlen J, Haanpää M, Hansson P, Jensen TS, Kemp H, Kennedy D, Leffler AS, Rice ASC, Segerdahl M, Serra J, Sindrup S, Solà R, Tölle T, Schuh-Hofer S, Treede RD, Pogatzki-Zahn E, Maier C, Vollert J. Pain thresholds and intensities of CRPS type I and neuropathic pain in respect to sex. Eur J Pain 2020;24:1058-1071.

van Rooijen DE, Marinus J, Schouten AC, Noldus LP, van Hilten JJ. Muscle hyperalgesia correlates with motor function in complex regional pain syndrome type 1. J Pain 2013;14:446-454.

Wan X, Wang W, Liu J, Tong T. Estimating the sample mean and standard deviation from the sample size, median, range and/or interquartile range. BMC Medical Research Methodology 2014;14:135.

Wittayer M, Dimova V, Birklein F, Schlereth T. Correlates and importance of neglect-like symptoms in complex regional pain syndrome. Pain 2018;159:978-986.

**<u>Supplement 4:</u> Risk of bias assessment of studies included in quantitative analysis**

Risk of bias

| Study | D1 | D2 | D3 | D4 | D5 | D6 | Overall |
|---|---|---|---|---|---|---|---|
| Louis et al (2025) | + | + | + | + | + | + | + |
| Schmidt et al (2025) | - | + | + | + | + | + | + |
| Schoenmacker et al (2023) | + | + | + | + | + | + | + |
| Habig et al (2021) | + | + | - | + | + | + | + |
| Dimova et al (2020) | + | + | + | - | + | + | + |
| Kinfe et al(2020) | + | - | - | + | + | + | + |
| Dietz et al (2019) | + | + | + | + | + | + | + |
| Meyer-FreBem et al(2019) | + | + | + | + | + | + | + |
| Konig et al (2019) | + | + | + | + | + | + | + |
| Wittayer et al (2018) | + | + | + | + | + | + | + |
| Kumowski et al (2017) | + | + | + | + | + | + | + |
| Enax-Krumova et al (2017) | + | + | + | + | + | + | + |
| Breuer et al (2014) | + | + | + | - | - | + | + |
| Reimer et al(2016) | + | + | + | + | + | + | + |
| Van Rooijen et al (2013) | + | + | + | - | + | + | + |
| Gierthmuhlen et al(2012) | + | + | + | + | + | + | + |
| Huge et al (2011) | + | + | + | + | - | + | + |
| Maier et al (2010) | + | + | + | + | + | + | + |
| Eberle et al(2009) | + | + | + | + | + | + | + |
| Huge et al (2008) | + | + | + | + | - | + | + |
| Uceyler et al 2007 | + | + | + | + | + | + | + |

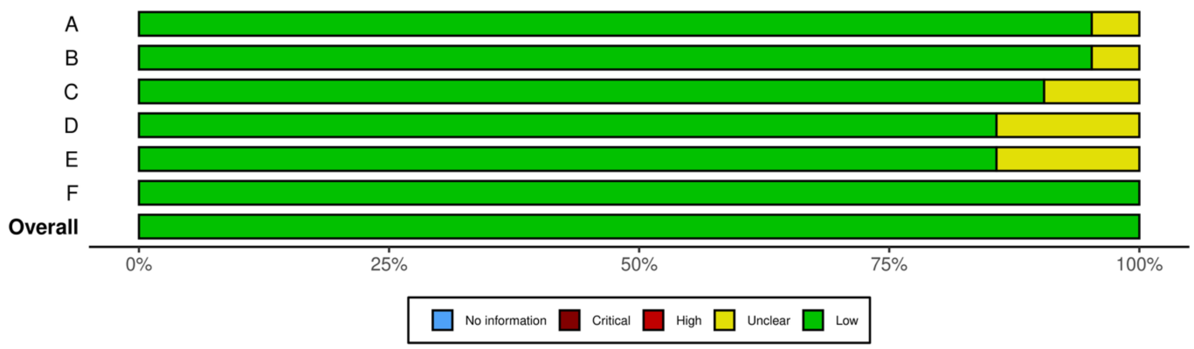


Traffic light plot and summary plot for risk of bias assessment of included studies. Green circles indicate low risk of bias, yellow circles indicate some concerns towards risk of bias. Risk of bias assessment included the following criteria:

**D1:** Was the research question or objective in this paper clearly stated?

**D2:** Was the study population clearly specified and designed?

**D3:** Were all the subjects selected or recruited from the same or similar populations (including the same time period)? Were inclusion and exclusion criteria prespecified and applied uniformly to all patients?

**D4:** Were QST parameters clearly defined and implemented consistently across all populations?

**D5:** Was a mechanism-based justification given for the use of individual QST parameter(s) based on underlying pathophysiological pain mechanisms?

**D6:** Were the outcome measures (independent variables) clearly defined, valid, reliable, and implemented consistently across all study participants?

**<u>Supplement 5:</u>** **Standard normal values (z-values) of QST parameters of all studies included in quantitative analysis (mean ± SE).**

**Part 1.** *Ipsilateral QST parameters of all studies <u>included</u> in analysis.* All values except DMA and PHS are provided as mean ± SEM of z-values. DMA is provided as logarithmic mean (0 was set 0.1) and standard error and PHS is provided as mean number of events ± SEM. n/a: data not available.

| **ipsilateral** | **n** | **CDT** | **WDT** | **TSL** | **CPT** | **HPT** | **PPT** | **MPT** | **MPS** | **WUR** | **MDT** | **VDT** | **DMA** | **PHS** |
|---|---|---|---|---|---|---|---|---|---|---|---|---|---|---|
| Breuer 2014 | 20 | n/a | n/a | n/a | n/a | 1.30 ± 0.30 | 1.88 ± 3.52 | n/a | n/a | n/a | n/a | n/a | n/a | n/a |
| de Schoenmaker 2023 | 21 | -1.00 ± 0.30 | -0.57 ± 0.45 | -1.13 ± 0.25 | 0.65 ± 0.25 | 0.80 ± 0.33 | 3.65 ± 0.48 | 1.75 ± 0.28 | 2.23 ± 0.33 | 0.60 ± 0.35 | -1.60 ± 0.35 | -1.58 ± 0.38 | -0.51 ± 0.51 | 0.30 ± 0.15 |
| Dietz 2019 | 105 | -1.03 ± 0.16 | -0.86 ± 0.14 | -0.93 ± 0.13 | 0.75 ± 0.12 | 0.72 ± 0.17 | 0.87 ± 0.34 | 1.15 ± 0.17 | 0.93 ± 0.16 | 0.13 ± 0.11 | 0.78 ± 0.18 | -1.33 ± 0.19 | n/a | n/a |
| Dimova 2020 | 202 | -1.28 ± 0.12 | -1.12 ± 0.10 | -1.26 ± 0.09 | 0.53 ± 0.10 | 0.58 ± 0.13 | 1.45 ± 0.43 | 1.02 ± 0.13 | 0.79 ± 0.12 | 0.05 ± 0.09 | 0.42 ± 0.15 | -1.64 ± 0.20 | -0.77 ± 0.04 | 0.26 ± 0.05 |
| Eberle 2009 | 50 | -1.74 ± 0.23 | -1.01 ± 0.20 | -1.46 ± 0.19 | 0.09 ± 0.19 | 0.63 ± 0.32 | 1.54 ± 0.25 | 0.47 ± 0.20 | 0.03 ± 0.19 | 0.45 ± 0.24 | -1.18 ± 0.34 | -3.28 ± 0.35 | -0.65 ± 0.09 | 0.73 ± 0.16 |
| Enax-Krumova 2017 | 24 | -0.50 ± 0.25 | -1.23 ± 0.28 | -0.90 ± 0.33 | 0.30 ± 0.30 | 0.13 ± 0.30 | 1.83 ± 0.30 | 0.90 ± 0.33 | 0.43 ± 0.33 | 0.68 ± 0.20 | -0.93 ± 0.40 | -0.48 ± 0.28 | n/a | n/a |
| Gierthmühlen 2012 | 344 | -1.05 ± 0.12 | -0.77 ± 0.11 | -0,77 ± 0.10 | 0.83 ± 0.10 | 1.14 ± 0.11 | 2.79 ± 0.17 | 0.61 ± 0.10 | 0.78 ± 0.11 | 0.37 ± 0.10 | -0.78 ± 0.13 | -1.45 ± 0.24 | -0.66 ± 0.51 | 0.13 ± 0.09 |
| Habig 2021 | 10 | -1.08 ± 0.26 | -1.58 ± 0.19 | -1.40 ± 0.14 | 1.62 ± 0.24 | 1.64 ± 0.49 | n/a | -0.15 ± 0.17 | 0.35 ± 0.25 | n/a | n/a | n/a | n/a | n/a |
| Huge 2008 (Acute) | 27 | -1.00 ± 0.30 | -1.38 ± 0.30 | -1.10 ± 0.30 | 1.08 ± 0.18 | 0.85 ± 0.28 | n/a | n/a | n/a | n/a | n/a | n/a | n/a | 1.26 ± 0.33 |
| Huge 2008 (Chronic) | 34 | -1.85 ± 0.25 | -1.65 ± 0.25 | -1.65 ± 0.18 | 0.63 ± 0.15 | 0.35 ± 0.40 | n/a | n/a | n/a | n/a | n/a | n/a | n/a | 0.12 ± 0.06 |
| Huge 2011 | 118 | -1.37 ± 0.14 | -1.42 ± 0.14 | -1.43 ± 0.13 | 1.01 ± 0.11 | 0.60 ± 0.14 | 2.81 ± 0.25 | 0.51 ± 0.11 | 0.66 ± 0.15 | 0.14 ± 0.09 | -1.53 ± 0.12 | -1.15 ± 0.18 | -0.72 ± 0.06 | 0.14 ± 0.50 |
| Kumovski 2017 | 24 | -0.83 ± 0.33 | -1.08 ± 0.35 | -1.15 ± 0.28 | 1.25 ± 0.20 | 0.78 ± 0.35 | 0.85 ± 1.00 | 0.23 ± 0.38 | 0.83 ± 0.35 | 0.55 ± 0.40 | 0.40 ± 0.33 | -0.65 ± 0.18 | -0.96 ± 0.62 | 0.18 ± 0.15 |
| König 2019 | 52 | -1.22 ± 0.25 | -0.87 ± 0.20 | -1.17 ± 0.22 | 0.73 ± 0.20 | 0.79 ± 0.28 | 1.32 ± 1.10 | 1.38 ± 0.25 | 0.98 ± 0.22 | -0.21 ± 0.17 | 0.62 ± 0.30 | -1.07 ± 0.49 | n/a | 0.11 ± 0.05 |
| Louis 2025 | 85 | -0.80 ± 0.12 | -0.75 ± 0.13 | -0.83 ± 0.10 | 1.13 ± 0.13 | 1.56 ± 0.18 | 1.78 ± 0.24 | 0.22 ± 0.13 | -0.30 ± 0.15 | 0.46 ± 0.18 | -1.15 ± 0.15 | -0.62 ± 0.21 | -0.85 ± 0.06 | 0.24 ± 0.07 |
| Maier 2010 | 403 | -1.07 ± 0.08 | -0.79 ± 0.08 | -0.90 ± 0.07 | 0.79 ± 0.06 | 1.06 ± 0.08 | 2.71 ± 0.15 | 0.46 ± 0.07 | 0.73 ± 0.08 | 0.31 ± 0.06 | -0.80 ± 0.09 | -1.54 ± 0.16 | -0.65 ± 0.03 | 0.23 ± 0.04 |
| Mainka 2013 | 18 | n/a | n/a | n/a | n/a | n/a | 2.39 ± 0.51 | n/a | n/a | n/a | n/a | n/a | n/a | n/a |
| Mehling 2024 | 20 | -1.62 ± 0.35 | -1.41 ± 0.31 | -1.65 ± 0.25 | 0.95 ± 0.19 | 0.83 ± 1.96 | 0.74 ± 0.32 | 1.98 ± 0.44 | 0.59 ± 0.25 | 0.13 ± 0.20 | -1.33 ± 0.46 | -2.94 ± 0.46 | n/a | n/a |
| Meyer-Frießem 2020 | 339 | -1.01 ± 0.13 | -0.79 ± 0.14 | -0.84 ± 0.12 | 0.79 ± 0.12 | 0.93 ± 0.19 | 2.60 ± 0.15 | 0.49 ± 0.18 | 0.80 ± 0.14 | 0.35 ± 0.12 | -0.67 ± 0.21 | -1.35 ± 0.27 | -0.78 ± 0.14 | 0.66 ± 0.12 |
| Reimer 2016 | 19 | -0.63 ± 0.33 | -0.95 ± 0.38 | -0.90 ± 0.35 | 0.88 ± 0.28 | 1.15 ± 0.48 | 3.33 ± 1.10 | 0.10 ± 0.20 | 0.53 ± 0.28 | 0.45 ± 0.20 | -0.98 ± 0.38 | -0.75 ± 0.50 | -0.68 ± 0.46 | 0.15 ± 0.15 |
| Schmidt 2024 | 11 | -0.97 ± 0.44 | -1.35 ± 0.43 | 0.30 ± 0.32 | n/a | 0.54 ± 0.43 | 2.94 ± 0.77 | 1.45 ± 0.41 | n/a | -0.63 ± 0.15 | -1.09 ± 0.57 | n/a | n/a | n/a |
| Üceyler 2007 | 32 | -1.70 ± 0.28 | -0.75 ± 0.18 | -0.95 ± 0.18 | 0.45 ± 0.20 | 0.23 ± 0.25 | 1.25 ± 0.23 | 0.50 ± 0.18 | 0.40 ± 0.25 | 0.40 ± 0.30 | -0.35 ± 0.48 | -1.65 ± 0.28 | -1.00 ± 0.00 | 0.65 ± 0.23 |
| Wittayer 2018 (Acute) | 20 | -0.80 ± 0.35 | -1.90 ± 0.40 | -0.90 ± 0.45 | 0.55 ± 0.35 | 0.10 ± 0.45 | 1.50 ± 1.15 | 1.10 ± 0.55 | 0.78 ± 0.40 | n/a | 0.05 ± 0.28 | -0.73 ± 0.40 | n/a | n/a |
| Wittayer 2018 (Chronic) | 33 | -1.65 ± 0.30 | -1.20 ± 0.30 | -1.40 ± 0.30 | 0.65 ± 0.30 | 1.00 ± 0.35 | 3.25 ± 1.55 | 1.18 ± 0.35 | 1.40 ± 0.35 | n/a | -0.45 ± 0.38 | -1.60 ± 0.50 | n/a | n/a |
| van Rooijen 2013 | 48 | -1.39 ± 0.32 | -1.65 ± 0.46 | n/a | 0.53 ± 0.18 | 0.30 ± 0.24 | 2.13 ± 0.22 | n/a | n/a | 0.25 ± 0.11 | n/a | -7.19 ± 2.17 | n/a | n/a |
| **Weighted mean ipsilateral (± SE)** | | **-1.13 ± 0.04** | **-0.95 ± 0.04** | **-0.99 ± 0.04** | **0.77 ± 0.04** | **0.89 ± 0.05** | **2.32 ± 0.08** | **0.66 ± 0.05** | **0.72 ± 0.05** | **0.28 ± 0.04** | **-0.56 ± 0.05** | **-1.42 ± 0.08** | **-0.72 ± 0.05** | **0.32 ± 0.04** |
| **n** | **2059** | **2013** | **2012** | **1963** | **2002** | **2032** | **1974** | **1899** | **1890** | **1797** | **1850** | **1926** | **1633** | **1742** |

**Part 2.** *Contralateral QST parameters of all studies included in quantitative analysis.* All values except DMA and PHS are provided as mean ± SEM of z-values. DMA is provided as logarithmic mean (0 was set to 0.1) and standard error and PHS is provided as mean number of events ± SEM. n/a: data not available.

| contralateral | n | CDT | WDT | TSL | CPT | HPT | PPT | MPT | MPS | WUR | MDT | VDT | DMA | PHS |
|---|---|---|---|---|---|---|---|---|---|---|---|---|---|---|
| de Schoenmaker 2023 | 21 | n/a | n/a | n/a | 0.18 ± 0.23 | 0.45 ± 0.25 | 1.88 ± 0.35 | 1.15 ± 0.23 | 1.38 ± 0.33 | 0.30 ± 0.33 | n/a | n/a | -1.00 ± 0.04 | n/a |
| Dietz 2019 | 105 | -0.48 ± 0.12 | -0.36 ± 0.11 | -0.45 ± 0.10 | 0.18 ± 0.10 | 0.29 ± 0.15 | -1.40 ± 0.13 | 0.83 ± 0.15 | 0.24 ± 0.14 | -0.22 ± 0.11 | 1.36 ± 0.11 | -1.15 ± 0.20 | n/a | n/a |
| Dimova 2020 | 202 | -0.56 ± 0.08 | -0.40 ± 0.09 | -0.52 ± 0.08 | -0.08 ± 0.09 | 0.25 ± 0.11 | -1.40 ± 0.10 | 0.83 ± 0.11 | 0.28 ± 0.10 | -0.28 ± 0.08 | 1.19 ± 0.10 | -1.11 ± 0.14 | -0.96 ± 0.02 | 0.17 ± 0.04 |
| Eberle 2009 | 50 | -0.42 ± 0.15 | -0.06 ± 0.16 | -0.41 ± 0.13 | -0.16 ± 0.13 | 0.51 ± 0.20 | 0.16 ± 0.16 | 0.69 ± 0.16 | -0.21 ± 0.18 | 0.32 ± 0.26 | 0.65 ± 0.18 | -2.03 ± 0.38 | -0.91 ± 0.04 | 0.30 ± 0.10 |
| Habig 2021 | 10 | -0.05 ± 0.17 | -0.44 ± 0.17 | -0.54 ± 0.14 | 0.60 ± 0.37 | 0.48 ± 0.45 | n/a | -0.10 ±0.11 | 0.40 ± 0.14 | n/a | n/a | n/a | n/a | n/a |
| Huge 2008 (Acute) | 27 | -0.23 ± 0.20 | -0.55 ± 0.23 | -0.43 ± 0.20 | 0.80 ± 0.18 | 1.00 ± 0.23 | n/a | n/a | n/a | n/a | n/a | n/a | n/a | 1.11 ± 0.30 |
| Huge 2008 (Chronic) | 34 | -1.40 ± 0.20 | -1.28 ± 0.20 | -1.05 ± 0.15 | 0.50 ± 0.20 | 0.28 ± 0.25 | n/a | n/a | n/a | n/a | n/a | n/a | n/a | 0.00 ± 0.00 |
| Huge 2011 | 118 | -0.92 ± 0.12 | -1.07 ± 0.13 | -1.02 ± 0.12 | 0.75 ± 0.11 | 0.32 ± 0.13 | 0.96 ± 0.17 | 0.33 ± 0.08 | 0.41 ± 0.14 | 0.18 ± 0.01 | -0.81 ± 0.10 | -1.23 ± 0.21 | -0.89 ± 0.06 | 0.11 ± 0.57 |
| König 2019 | 52 | -0.33 ± 0.14 | -0.40 ± 0.18 | -0.49 ± 0.16 | 0.34 ± 0.17 | 0.46 ± 0.24 | -1.23 ± 0.34 | 1.46 ± 0.17 | 0.72 ± 0.17 | -0.60 ± 0.12 | 1.71 ± 0.10 | -0.53 ± 0.20 | n/a | 0.13 ± 0.05 |
| Louis 2025 | 85 | -0.30 ± 0.11 | -0.33 ± 0.11 | -0.40 ± 0.09 | 1.00 ± 0.12 | 1.87 ± 0.15 | 0.44 ± 0.16 | 0.09 ± 0.12 | -0.76 ± 0.12 | 0.20 ± 0.18 | -0.76 ± 0.12 | -0.88 ± 0.14 | -1.00 ± 0.00 | n/a |
| Maier 2010 | 403 | -0.49 ± 0.06 | -0.22 ± 0.06 | -0.34 ± 0.06 | 0.40 ± 0.05 | 0.58 ± 0.10 | 0.29 ± 0.10 | 0.05 ± 0.06 | 0.11 ± 0.06 | 0.09 ± 0.05 | -0.09 ± 0.07 | -0.82 ± 0.10 | -0.93 ± 0.01 | 0.17 ± 0.03 |
| Mehling 2024 | 20 | -1.37 ± 0.22 | -0.57 ± 0.25 | -0.46 ± 0.24 | 0.41 ± 0.16 | -0.22 ± 0.20 | -0.06 ± 0.31 | 0.91 ± 0.34 | 0.61 ± 0.28 | 0.00 ± 0.16 | -0.83 ± 0.26 | -1.94 ± 0.33 | n/a | n/a |
| Reimer 2016 | 19 | 0.15 ± 0.23 | -0.28 ± 0.33 | -0.30 ± 0.18 | 0.75 ± 0.25 | 0.08 ± 0.23 | 1.20 ± 1.35 | 0.05 ± 0.20 | 0.00 ± 0.28 | 0.60 ± 0.20 | -0.10 ± 0.23 | -0.63 ± 0.55 | -1.00 ± 0.00 | 0.00 ± 0.00 |
| van Rooijen 2013 | 48 | -0.35 ± 0.27 | -0.52 ± 0.24 | n/a | 0.56 ± 0.17 | 0.41 ± 0.17 | 1.18 ± 0.15 | n/a | n/a | n/a | n/a | -2.15 ± 0.81 | n/a | n/a |
| **Weighted mean contralateral (± SE)** | | **-0.54 ± 0.03** | **-0.42 ± 0.04** | **-0.49 ± 0.03** | **0.37 ± 0.03** | **0.52 ± 0.05** | **-0.08 ± 0.05** | **0.43 ± 0.04** | **0.17 ± 0.04** | **0.00 ± 0.03** | **0.27 ± 0.04** | **-1.07 ± 0.06** | **-0.94 ± 0.01** | **0.18 ± 0.04** |
| **n** | **1194** | **1165** | **1165** | **1117** | **1186** | **1186** | **1115** | **1074** | **1076** | **1024** | **1044** | **1092** | **894** | **900** |

**<u>Supplement 6:</u> Ipsilateral and contralateral standard normal values (z-values) of QST parameters stratified for CRPS duration across all studies included in quantitative analysis**

| | Ipsilateral (affected site) | | | contralateral | | |
|---|---|---|---|---|---|---|
| | **early** | **intermediate** | **late** | **early** | **intermediate** | **late** |
| **CDT** | -1.06 ± 0.08 | -1.11 ± 0.06 | -1.17 ± 0.09 | -0.27 ± 0.08 | -0.49 ± 0.04 | -1.03 ± 0.10 |
| **WDT** | -1.02 ± 0.09 | -0.88 ± 0.05 | -0.97 ± 0.09 | -0.28 ± 0.08 | -0.30 ± 0.04 | -1.12 ± 0.11 |
| **TSL** | -1.03 ± 0.08 | -0.98 ± 0.05 | -0.97 ± 0.09 | -0.40 ± 0.07 | -0.40 ± 0.04 | -1.03 ± 0.10 |
| **CPT** | 0.74 ± 0.07 | 0.74 ± 0.05 | 0.85 ± 0.09 | 0.62 ± 0.08 | 0.24 ± 0.04 | 0.63 ± 0.09 |
| **HPT** | 0.85 ± 0.10 | 0.90 ± 0.07 | 0.94 ± 0.10 | 1.17 ± 0.10 | 0.44 ± 0.07 | 0.33 ± 0.10 |
| **PPT** | 1.68 ± 0.16 | 2.24 ± 0.12 | 2.84 ± 0.12 | 0.44 ± 0.15 | -0.49 ± 0.07 | 1.10 ± 0.15 |
| **MPT** | 0.43 ± 0.09 | 0.68 ± 0.07 | 0.65 ± 0.09 | 0.29 ± 0.10 | 0.45 ± 0.05 | 0.45 ± 0.08 |
| **MPS** | 0.18 ± 0.09 | 0.81 ± 0.06 | 0.81 ± 0.10 | -0.44 ± 0.10 | 0.22 ± 0.05 | 0.56 ± 0.13 |
| **WUR** | 0.48 ± 0.10 | 0.24 ± 0.05 | 0.30 ± 0.09 | 0.31 ± 0.13 | -0.09 ± 0.04 | 0.19 ± 0.03 |
| **MDT** | -0.77 ± 0.11 | -0.33 ± 0.07 | -1.02 ± 0.10 | -0.20 ± 0.09 | 0.56 ± 0.05 | -0.81 ± 0.10 |
| **VDT** | -1.29 ± 0.11 | -1.46 ± 0.11 | -1.38 ± 0.14 | -1.23 ± 0.15 | -0.92 ± 0.07 | -1.23 ± 0.21 |
| **DMA** | -0.82 ± 0.06 | -0.72 ± 0.07 | -0.67 ± 0.16 | -0.97 ± 0.01 | -0.94 ± 0.01 | -0.90 ± 0.05 |
| **PHS** | 0.51 ± 0.06 | 0.38 ± 0.04 | 0.14 ± 0.21 | 0.47 ± 0.08 | 0.17 ± 0.02 | 0.09 ± 0.39 |

All parameters except DMA and PHS are displayed as weighted mean ± SEM of z-values. DMA is presented as logarithmic mean ± SEM (-1.00 representing 0 pain rating). PHS is presented as mean number of events ± SEM.

**<u>Supplement 7:</u> QST as an analytical tool for response to therapy**

Five studies, in aggregate only comprising 57 patients, used the DFNS type of QST profiling to investigate the impact of therapeutic intervention. A three-day treatment with the NSAID parecoxib in 10 patients did not significantly impact on the QST profile (Breuer et al. 2014). The pain-modulating capacities of C-tactile fibre (affective touch) stimulation was assessed in the hairy skin in 10 CRPS patients (Habig et al. 2021). They reported a normalisation of thermal pain thresholds (both cold and heat pain), while thermal detection thresholds showed persistent sensory loss, PPT was not assessed. However, pain intensity in CRPS was not reduced by repeated C-tactile-targeted touch.

A case report showed that treatment with low-dose anti-TNF alpha antibody resulted in approximately 50% reduction selective for pressure hyperalgesia, but not other pain parameters and almost complete elimination of dynamic mechanical allodynia (Bernateck et al. 2007).

The impact of multimodal pain treatment was tested in 24 patients before and at 6 months follow-up (Enax-Krumova et al. 2017). The only QST parameter with a significant number of abnormal pain sensitivity findings at baseline was PPT and pain relief was associated with its normalisation. Abnormally sensitized PPT was encountered in 12/24 patients (50%). A relevant treatment response at follow-up was defined as pain relief > 30% and reached by 17/24 patients. This subgroup reported 7/17 (41%) of abnormal PPT assessments in the treatment responder group, which dropped to 2/17 = 12% at follow-up (p=0.052), but not in non-responders (5/7 = 72% to 4/7 = 57%, p=0.58; Enax-Krumova et al. 2017).

A cohort of 12 patients with longstanding CRPS of the knee was tested before and at 3 months follow-up after unilateral L4 dorsal root ganglion (DRG) stimulation (Kinfe et al. 2020). The contralateral QST was used as a reference. L4-DRG stimulation reduced sensory gain in CDT, PPT and WUR and reduced sensory loss in MPT and MPS, while detection parameters were not altered at follow-up. Notably, L4-DRG stimulation reduced pressure pain sensitivity significantly to the level of the contralateral unaffected knee. Pain relief was significantly associated with normalization of pain QST (r=0.51, p<0.05). Half of the patients reported a substantial average pain relief (− 60±11%, p<0.005), which paralleled substantial QST normalisation, but neither pain reduction nor QST normalisation was found in the non-responder subgroup (Kinfe et al. 2020).

<u>References:</u>

Bernateck M, Rolke R, Birklein F, Treede RD, Fink M, Karst M. Successful intravenous regional block with low-dose tumor necrosis factor-alpha antibody infliximab for treatment of complex regional pain syndrome 1. *Anesth Analg*. 2007;105:1148-51

Breuer AJ, Mainka T, Hansel,N, Maier C, and Krumova, EK. Short-term treatment with parecoxib for complex regional pain syndrome: a randomized, placebo-controlled double-blind trial. Pain Physician 2014;17:127-137.

Enax-Krumova EK, Lenz M, Frettloh J, et al. Changes of the Sensory Abnormalities and Cortical Excitability in Patients with Complex Regional Pain Syndrome of the Upper Extremity After 6 Months of Multimodal Treatment. Pain Med 2017;18:95-106.

Habig, K., Lautenschlager, G., Maxeiner, H., Birklein, F., Kramer, H.H., and Seddigh, S. Low mechano-afferent fibers reduce thermal pain but not pain intensity in CRPS. BMC Neurol 2021;21:272.

Kinfe T, von Willebrand N, Stadlbauer A, Buchfelder M, Yearwood TL, Muhammad S, Chaudhry SR, Gravius S, Randau T, Winder K, Maihöfner C, Gravius N, Magerl W. Quantitative sensory phenotyping in chronic neuropathic pain patients treated with unilateral L4-dorsal root ganglion stimulation. J Transl Med. 2020;18:403
